# Refining the genetic landscape of anophthalmia and microphthalmia: a comprehensive framework with deep learning and updated gene panels

**DOI:** 10.1101/2025.08.26.25334245

**Authors:** Mara I. Maftei, Lucas G.N. Spink, Oriol Gracia Carmona, Simona Mikula Mrstakova, Lean Abahreh, Robin Hayes, Aitor Bañon, María Elisa Cuevas, Katherine Cid, Raul Araya-Secchi, Franca Fraternali, Jing Yu, Gavin Arno, Rodrigo M. Young

## Abstract

**Importance:** Anophthalmia and microphthalmia (A/M) are rare congenital eye disorders with a low molecular diagnosis rate, which limits clinical management and genetic counselling. Improved detection and interpretation of pathogenic variants is essential for advancing diagnosis and care in affected individuals.

**Objective:** To improve the molecular diagnostic yield in A/M patients by refining the methodology of variant investigation and association using an updated rigorously curated gene panel, and a refined bioinformatic pipeline incorporating structural variant detection, *in silico* Artificial Intelligence assisted predictive tools, and molecular dynamics simulations.

**Methodology:** We curated an updated A/M gene panel through a systematic literature review and screened for rare variants in these genes using data from the UK’s 100,000 Genomes Project, a national whole-genome sequencing initiative conducted by Genomics England. The cohort comprised 306 individuals recruited to the Rare Disease programme with a clinical diagnosis of anophthalmia or microphthalmia, recorded either as the primary phenotype or within HPO, SNOMED, or ICD-10 terms. Variants, including loss-of-function, missense, RNA splicing, and structural variants, were annotated with deep learning tools (AlphaMissense, SpliceAI), and missense variants were further assessed using REVEL, Missense3D, and molecular dynamics simulations.

**Results:** We identified pathogenic or likely pathogenic variants in 37 (12.1%) individuals, with an additional 23 (7.5%) harbouring strong candidate variants of uncertain significance. Our literature review identified the biggest contributors to A/M phenotypes to be *MFRP*, *OTX2*, *PRSS56* and *SOX2*, each with over 100 patients reported in the literature, with a total number of 124 genes found be associated to A/M. Variants from our screen were most often found in genes with high A/M association, but also included novel findings within genes with a weaker association to A/M such as *ACTG1*, *HDAC6*, *RERE* and *SIX3;* adding support to their disease relevance.

**Conclusions and Relevance:** This study increased the diagnostic yield in A/M patients recruited to the 100KGP, and provides further evidence of genotype-phenotype associations within the aetiology of A/M. We also provide an updated framework for enhancing clinical genetic diagnosis in A/M that may inform broader strategies for other complex congenital disorders. However, as molecular diagnosis of A/M remains low, further research in understanding the genetic aetiology of A/M is necessary.

## Introduction

Cost-effective, high-throughput whole genome sequencing has significantly improved our understanding of the genetic basis of rare diseases^1,2^. However, many cases remain unsolved, limiting genetic counselling and patient care^3,4^. In particular, the molecular diagnosis rate in anophthalmia (A; absence of the eye) and microphthalmia (M; small eye) is only 13–33%^5–14^. Although A/M is untreatable, identifying a genetic cause remains crucial to enable appropriate clinical management, monitoring for associated complications, and enabling cascade testing and family reproductive planning^12,15^.

A/M constitutes a spectrum of disease that can present unilaterally or bilaterally; occur in isolation or alongside ocular anomalies such as coloboma or anterior segment dysgenesis^11,12^; and often form part of syndromic presentations with other systemic abnormalities^16^. More than 100 genes have been associated to A/M, with the literature reporting *SOX2* as the most frequently disrupted gene (15–40% of diagnosed cases), followed by *OTX2* and *RAX*^10,11,14^. Methodological limitations and knowledge gaps may contribute to the low molecular diagnosis rate of A/M. Clinical and genetic heterogeneity, such as incomplete penetrance, variable expressivity, and mosaicism can complicate variant interpretation for molecular diagnosis^11,12^. Furthermore, underlying environmental factors, such as foetal alcohol syndrome, maternal vitamin A deficiency, exposure to toxins and certain infections during gestation may also impact the outcome of A/M without a monogenic cause^17^.

The 100,000 Genomes Project (100KGP)^18^ has enabled the systematic aggregation of genomic and clinical data of rare disease patients in the UK, providing an opportunity to investigate genetically heterogeneous conditions such as A/M in a phenotypically profiled cohort^1^. To address the persistently low molecular diagnosis rate of A/M, we set out to implement a new genomic variant interrogation pipeline to enable comprehensive evaluation of all A/M participants within the 100KGP cohort. This approach involved curating an A/M gene panel based on a systematic literature review, incorporating SV discovery, and utilising Alphamissense and SpliceAI, deep learning-based *in silico predictors,* to interpret variant pathogenicity. Our strategy led to the identification of 60 candidate variants in 56 previously unsolved participants, thereby strengthening the genotype–phenotype correlations for *ACTG1*, *HDAC6*, *RERE* and *SIX3* which are less commonly associated with A/M.

## Methods

### A/M Cohort Selection

Participants with a diagnosis of A/M were identified in the 100KGP dataset using relevant Human Phenotype Ontology (HPO) terms, ICD-10 codes, and/or SNOMED terms. This approach yielded a cohort of 306 individuals from 283 unrelated families. Notably, 118 participants from 108 families were recruited with A/M as the disease term, while the remaining 188 participants had A/M included in their HPO terms or clinical (SNOMED or ICD-10) terms, but were recruited under a different clinical condition. Of note, in compliance with Genomics England Airlock policy, detailed clinical terms are not included herein^18^.

### A/M Gene Panel Curation

A 124 A/M-associated gene panel (eTable1) was generated based on the Genomics England PanelApp ‘Green’ and ‘Amber’ genes (2016)^19^, and literature reviews: Patel et al. (2018)^10^, Harding and Moosajee (2019)^11^, and Plaisancié et al. (2019)^12^. Additional genes identified in recent studies were included: *EPHA2*^13,20,21^, *GJA8*^1,32–38^, *NR6A1*^22^, and *PRR12*^23,24^. We systematically reviewed published reports of variants in genes included in the panel, using PubMed keyword searches “gene”, “human”, “microphthalmia” or “anophthalmia”. Only individuals harbouring a variant and explicitly described to have anophthalmia, microphthalmia, or documented axial length measurements consistent with an A/M diagnosis were included. Genes having pathogenic variants reported in at least five individuals across two or more families were defined as those with the strongest evidence for an association.

### Genomic Variant Calling and Annotation

Structural variants were identified using the SVRare pipeline^25^, with genomic coordinates aligned to the GRCh38 (hg38) assembly. Small variants were filtered for rarity, with an allele frequency (AF) of ≤0.001 in gnomAD v3.25^26^ and the Genomics England research cohort, with retention of missense and splice-altering variants meeting in silico deleteriousness thresholds (AlphaMissense >0.340; SpliceAI >0.5). (see Supplementary Methods). To avoid artifacts or miscalls, all candidate variants were confirmed by inspection using the Integrative Genomics Viewer (IGV^27^), with a minimum read depth of 15x and mapping quality ≥ 10. Subsequently, all true missense calls were annotated manually with REVEL^28^ and Missense3D^29^. Furthermore, all missense variants located within functional domains were subjected to molecular dynamics (MD) simulations through a Root Mean Square Fluctuation analysis (see Supplementary Methods).

## Results

### Candidate variant identification

Sixty variants (in 56 individuals, eTable2, Fig.S1) within the A/M gene panel passed our filtering criteria: 16 pLOF (predicted loss-of-function), 7 structural variants (SV), 6 candidate splice-disrupting variants and 31 missense variants (summarised in eTable2). 37 participants (eTable2; 12.1% of the A/M cohort) harboured pathogenic or likely pathogenic variants and 23 (7.5%) carried candidate rare variants (AF ≤0.001) of unknown significance (VUS) with high pathogenicity scores. We consider 8 of these VUS (*ABCB6*:p.Met421Ile and p.Glu446Gly; *ALDH1A3*:p.Phe35Ser; *CHD7*:p.His1682Arg; *CRYBB2*:p.Arg145Pro; *NHS*:c.565+103674G>T; *OTX2*:p.Lys100Glu; and *TFAP2A*:p.Ala299Asp) to be likely pathogenic based on their location within critical functional domains, consistently high in silico pathogenicity predictions, and/or their strong specificity to the observed phenotypes. Participants 15, 43 and 46 carried candidate compound heterozygous variants in *ERCC2, PRSS56* and *RAB3GAP1*, respectively (eTable2).

### Predicted loss-of-function alleles

All participants harbouring pLOF variants had phenotypes, inheritance and segregation patterns consistent with the clinical presentations associated with these gene defects (eTables 2 and 3). The variants *KMT2D:*p.Met1379ValfsTer52 (participant 23) and *SOX2:*p.Asn24ArgfsTer65 (participant 53) had both been reported in unrelated cases of A/M^30–36^.

A recurrent *PAX2* frameshift variant (p.Val26GlyfsTer28) was identified in participant 35. This variant was also present in six other 100KGP participants, five with renal-coloboma syndrome features without microphthalmia, and one unaffected relative. This variant has been well documented as pathogenic and has variable expressivity including kidney hypoplasia and ocular involvement^37–39^.

### Possible mosaicism in A/M genes

Three pLoF variants were observed affecting 10-20% of WGS reads, suggesting possible mosaicism: *BCOR*:p.Ser286AlafsTer92 (21.4% affected reads, participant 6, Fig.S1A), *BCOR*:p.Val1286CysfsTer8 (11.5% affected reads, participant 8, Fig.S1B), and *MYRF*:p.Pro190ArgfsTer19 (18.5% affected reads, participant 26, Fig.S13). Participant 26 expressed phenotypes milder than what is expected for a *MYRF* LOF variant, possibly due to mosaicism^40–42^ (eTable2), while the participants 6 and 8 have clinical phenotypes (eTable2) in keeping with the known phenotypic variability in mosaic *BCOR* cases^43,44^.

### Structural variant analysis

Six SVs impacting the coding sequence of *ATOH7*, *HCCS*, *NDP*, *NHS*, *OTX2* and *PTCH1* were identified and classified as pathogenic or likely pathogenic according to ACMG/ClinGen guidelines^45^ (eTable2).

Large deletions encompassing entire genes within our panel were identified in participants 5, 18 and 53 (eTable2). Participant 5 carries a 14.6 Mb (10:57,171,179-71,785,816) heterozygous deletion (Fig.S2A) spanning 72 coding genes including *ATOH7*, *ANK3*, *NEUROG3*, and *NODAL*, all involved in early embryonic development. Participant 18 and her affected mother (18_M) carry a 471.5 kb (X:10,945,466-11,417,014) deletion encompassing *HCCS* (Fig.S2B), and three other coding genes: *ARHGAP6, AMELX, MIR548AX*. Participant 53 harbours a *de novo* 707.7 kb (14:56,783,923-57,491,668; Fig.S2C) deletion deleting the entire *OTX2*, and *EXOC5, AP5M1, NAA30, CCDC198*.

SVs partially deleting genes were identified in participants 28, 29 and 41 (Fig.S2D-F, eTable2). Participant 28 inherited a hemizygous 4.4 kb (X:43,954,691-43,959,091) deletion from his heterozygous, unaffected mother. This variant resulted in the deletion of exon 2 in *NDP*, leading to start codon loss (Fig.2A, Fig.S2D). Participant 29 inherited a hemizygous 44.5 kb (X:17,153,721-17,599,035) deletion of *NHS* exon 1 from his mother (29_M) who expresses similar, but milder phenotypes (Fig.2B, Fig.S2E, eTable 3). Finally, a *de novo* 1.8 kb (9:95,475,579-95,477,447) deletion of *PTCH1* exons 11–12 was identified in participant 44 (Fig.1Y, Fig.S2F).

**Figure 1.**
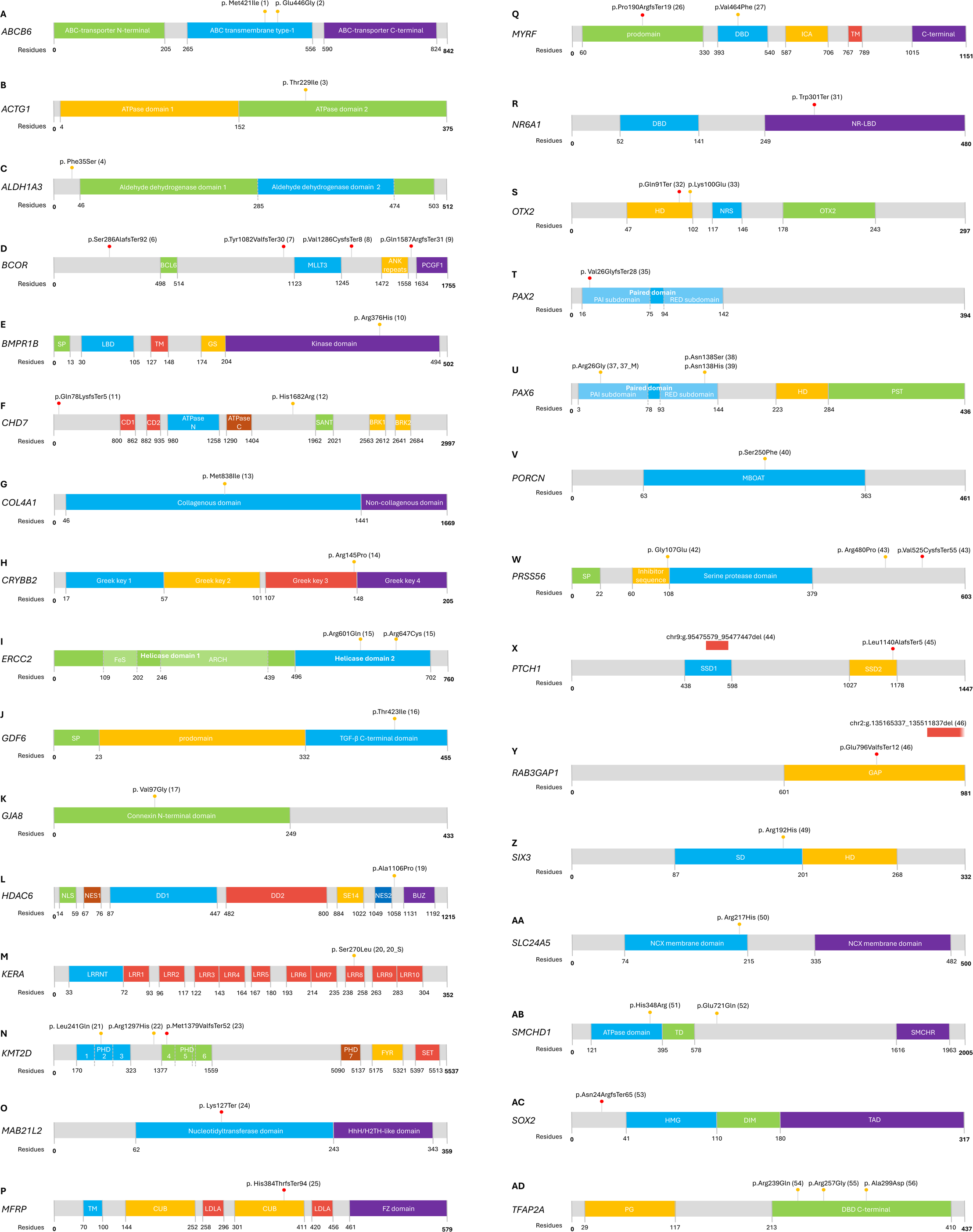
Schematic of coding variants found within A/M participants of the 100K GP. A-AD) Schematic of the coding variants identified by our screening within anophthalmic and microphthalmic patients recruited to the 100KGP. Predicted-loss-of-function variants are represented in red and missense in yellow. The participants who harbour each variant is presented in parentheses.

**Figure 2.**
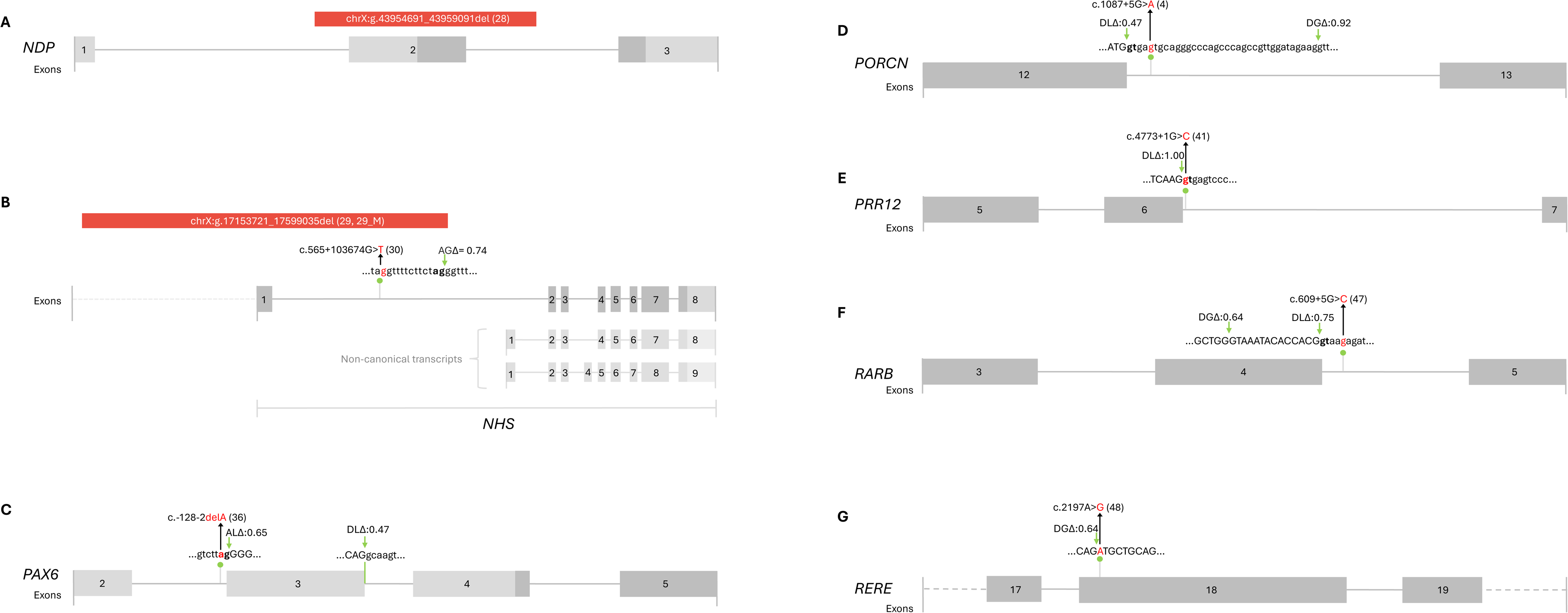
Schematic of variants affecting non-coding regions found within A/M participants of the 100KGP. A-H) Schematic of the variants affecting non-coding regions that were identified by our screening within anophthalmic and microphthalmic patients recruited to the 100KGP. Splice variants are represented in green, with the participants who harbour each variant presented in parentheses. AG: acceptor gain, AL: acceptor loss, DG: donor gain, DL: donor loss.

### Identification of candidate RNA splice-altering variants

Six variants that potentially disrupt RNA splicing were identified: *NHS*:c.565+103674G>T (Fig.2B, participant 30), *PAX6*:c.-128-2del (Fig.2C, participant 36), *PORCN*:c.1087+5G>A (Fig.2D, participant 4), *PRR12*:c.4773+1G>C (Fig.2E, participant 41), *RARB*:c.609+5G>C (Fig.2F, participant 47), *RERE*:*c.2197A>G* (Fig.2G, participant 48). The variants in *PAX6* and *PRR12* carried by participants 36 and 41 were predicted to be pathogenic, while the variants in *NHS*, *PORCN*, *RARB* and *RERE* are classified as VUS.

The *NHS*:c.565+103674G>T and *PORCN*:c.1087+5G>A variants were both identified hemizygously in male participants (30 and 4, respectively), inherited from their unaffected heterozygous mothers (eTable 5). For the *NHS*:c.565+103674G>T variant, SpliceAI predicts an acceptor site gain (Δscore=0.74) 13bp downstream of the variant position (Fig.2B). This could lead to pseudo-exon inclusion in the transcript. Notably, this variant was also identified within another 100KGP participant with HPO terms in keeping with Nance Horan Syndrome, but not affected with microphthalmia. Functional investigation of the splice impact would clarify its pathogenicity in these cases.

The *PORCN* variant, located at the conserved +5 position of intron 12, showed a SpliceAI donor loss score of 0.47 and a high-confidence alternative donor gain site 31bp downstream (Δscore=0.92, Fig.2D). Use of this alternate donor gain would result in a 36bp intronic inclusion and an in-frame insertion (p.His362_Val363ins12) resulting in a disruption of the MBOAT domain, potentially disrupting Wnt signalling^46^.

Heterozygous variants in *RARB*:c.609+5G>C and *RERE*:c.2197A>G (p.Met733Val) were identified in participants 47 and 48, respectively. The first, in intron 4 is predicted to abolish the canonical donor site (Δscore=0.75) and activate a cryptic donor site 19bp upstream (Δscore=0.64), leading to deletion of 14bp of exon 4 from the transcript, and resulting in a predicted frameshift p.Gly199GlufsTer3 (Fig.2F). The *RERE* missense variant (p.Met733Val), while scoring low on missense pathogenicity prediction (AlphaMissense score 0.069), is predicted to create a strong cryptic splice donor site within exon 18 (Δscore=0.97). This would lead to the excision of 1,199 nucleotides, causing a large deletion and frameshift (p.Met733ValfsTer35 Fig.2G).

### Missense variant analysis

We identified thirteen heterozygous missense variants in genes associated with autosomal dominant A/M, two variants in genes associated with X-linked A/M, two homozygous variants in recessive A/M genes and two compound heterozygous genotypes (eTable2). Of these 31 missense variants, 15 were classified as pathogenic or likely pathogenic with the remaining as VUS.

All VUS were predicted pathogenic by AlphaMissense, with corresponding high REVEL scores (eTable2). Noteworthy, dominant *CHD7*, *GDF6*, *KMT2D* and *SMCHD1* variants were inherited from apparently unaffected parents (eTable2, eTable3) either suggesting these are not pathogenic or exhibit incomplete/non penetrance^10–12,47–49^. The VUSs in *ABCB6*, *COL4A1*, *CRYBB2*, *GDF6*, *KERA*, *KMT2D* (p.Leu241Gln) *MYRF, OTX2*, *PRSS56* (p.Gly107Glu), *SMCHD1* and *TFAP2A* impacted functional domains: ABC transporter transmembrane domain (*ABCB6*, Fig.1A), a collagenous domain (*COL4A1*, Fig.1G), a ‘Greek key’ domain (*CRYBB2*, Fig.1H), a TGF-β domain (*GDF6*, Fig.1J), a Leucine-rich repeat (*KERA*, Fig.1M), a PHD finger (*KMT2D*, Fig.1N), DNA binding domains (*MYRF*, Fig.1Q; *OTX2*, Fig.1S; *TFAP2A*, Fig.1AD) and a serine protease inhibitor domain (*PRSS56*, Fig1.W). While not affecting functional domains, the VUS variants in *ALDH1A3*, *CHD7*, *KMT2D* and *SMCHD1* (p.Glu721Gln), were carried by participants expressing phenotypes in keeping with with those reported for similar variants in these genes^48,50,51^. Notably, the homozygous p.Phe35Ser variant in *ALDH1A*3, was found in the same participant who carried the heterozygous *PORCN*:c.1087+5G>A variant. As both are VUS, further investigation is necessary, including RNA analysis for a splice defect in *PORCN,* to help determine which is more likely to be causative^52–54^.

Molecular dynamic simulations of protein domains can be used to study the effect of amino acid changes by analysing the Root Mean Square Fluctuation (RMSF) of atoms^55,56^. To add evidence to the pathogenicity of the identified missense VUS, we performed RMSF analyses on the missense variants within functional domains in *GDF6, MYRF, OTX2, PAX6, PORCN, SIX3, TFAP2A*, focusing on the Cα atoms to assess potential impacts on structural stability (Fig.3A). Comparisons centred on the top 10% of residues exhibiting the greatest fluctuation differences. The *PAX6* variants (p.Arg26Gly and p.Asn138His) displayed a clear destabilising effect on the whole modelled protein region (Fig.3A-C). *TFAP2A*:Arg239Gln also displayed a significant destabilising effect (Fig.3A, D). In contrast, the *SIX3*:p.Arg192His variant demonstrated a stabilising effect, suggesting increased conformational rigidity in the protein fold (Fig.2A, E).

**Figure 3.**
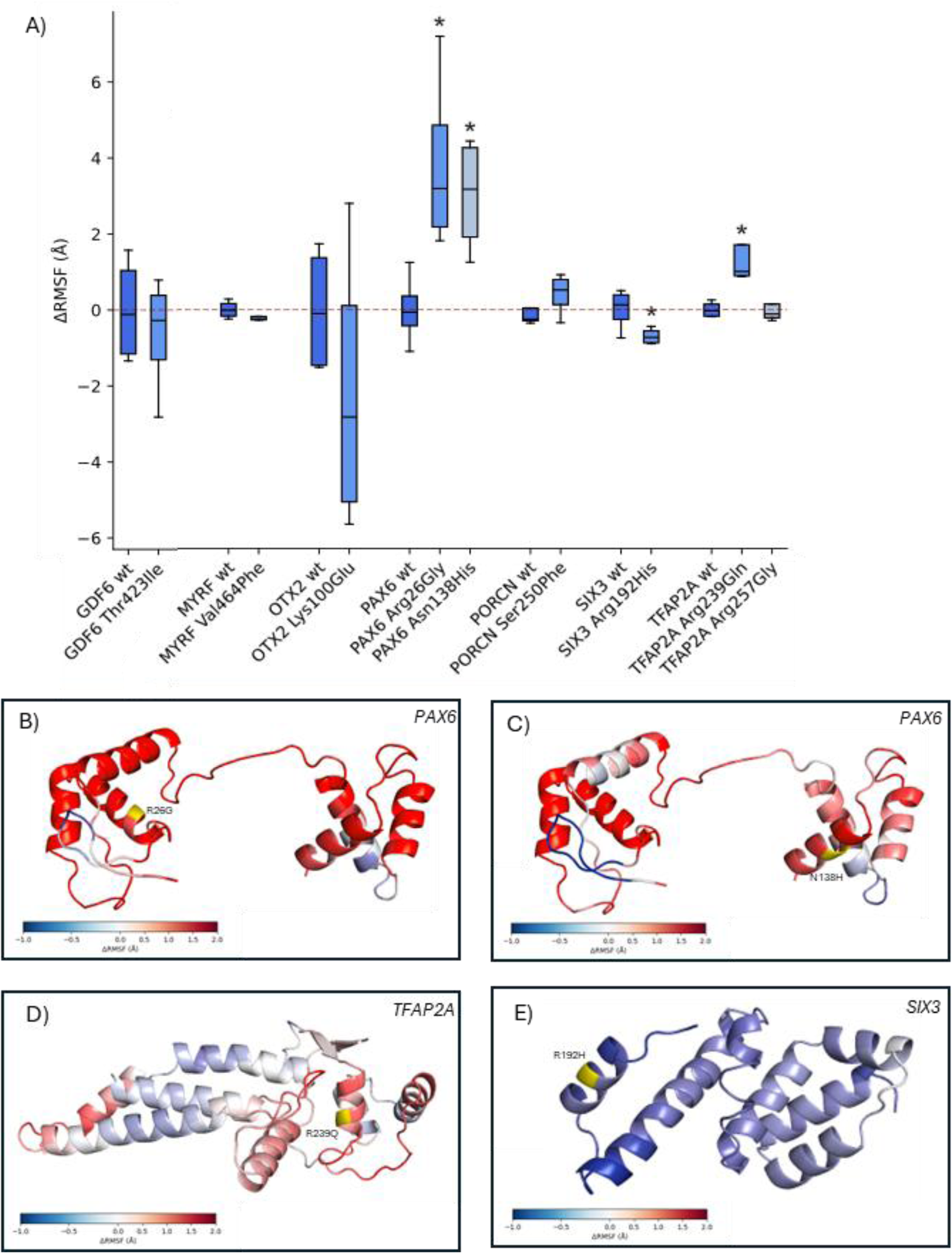
Root Mean Square Fluctuation for missense variants. A) Box plot showing Root Mean Square Fluctuation (RMSF) differences (in Å) for backbone Cα atoms, relative to the wild-type (wt) mean RMSF for each system. Differences were calculated using the top 10% of residues with the highest fluctuation differences in each system. Asterisks indicate statistical significance: p < 0.05(*). B-E) Protein structural representation of RMSF for those which achieved statistical significance in (A). Regions in red represent mutant flexibility compared to wt, while blue represents rigidity. Yellow represents the position of the amino acid substitution.

### Compound heterozygous variants

Participant 15 harboured two missense variants in *ERCC2* (Fig.1I, eTable2). The maternally inherited allele harboured c.1802G>A, p.Arg601Gln, with c.1939C>T, p.Arg647Cys presumed in trans (paternal DNA unavailable; eTable3). Participant 46 carried a maternally inherited frameshift variant in *RAB3GAP1* (p.Glu796ValfsTer12) and a large paternally inherited 346.5kb deletion (2:135,165,337-135,511,837; Fig.S2G) encompassing part of *RAB3GAP1* and *ZRANB3,* resulting in the loss of the final exon of *RAB3GAP1* (Fig.1Y, Fig.S2G, eTable2). Participant 43 carried a likely pathogenic frameshift p.Val525CysfsTer55 in *PRSS56* on the paternal allele, and a VUS missense p.Arg480Pro variant on the maternal allele (Fig.1W, eTable2).

## Discussion

This study describes the genetic findings in 60/306 individuals recruited to the UK 100KGP for a curated 124 gene panel implicated in A/M. Our analysis using updated variant interpretation tools yielded an overall candidate variant identification rate of 19.6%. However, the overall diagnostic rate for A/M is likely higher due to prescreening for variants within *OTX2* and *SOX2* prior to recruitment to the 100KGP, these genes together account for approximately 20% of A/M cases^11,12,56–59^. The estimated diagnostic rate is projected to approximately 35.7%.

Importantly, our analysis included all participants recruited with A/M regardless of solved status; 12 variants (from 12 participants) identified here, had been previously reported and published. One additional previously reported variant, *PDGFRA*:p.Thr432Met was not retained in our final candidate list, as *PDGFRA* currently lacks sufficient evidence for association with A/M and was excluded from our curated gene panel.

From our literature search to generate a curated A/M gene panel, we better characterised the genetic aetiology of A/M. Historically, *SOX2*, *OTX2*, and *RAX* were considered the top three contributors to A/M^10,11^; however, we identified that *SOX2*, *PRSS56*, *MFRP*, and *OTX2* were the most frequent contributors to A/M, each with over 100 patients reported. It may also be important to note that variants in both *MFRP* and *PRSS56* cause microphthalmia strictly affecting the posterior segment. As expected, the majority of pathogenic variants identified in the screening occurred in genes with strong associations to A/M such as *BCOR*, *OTX2*, *PAX6* and *TFAP2A.* Our findings in *ERCC2* and *NDP* further reinforce their association to A/M, as both previously had five reported cases^61–69^. We also identified a likely pathogenic variant in *HDAC6* and a strong VUS candidate in *ACTG1*, both genes with limited evidence of association with A/M; where the association is supported by either a single previously reported affected individual, or where the association has only been inferred^70–73^. We further identified a likely pathogenic variant in *SIX3* (previously three A/M cases reported, eTable1) and *RERE* (two cases reported, eTable1), adding further support for the association of these genes to A/M.

This study identified seven structural variants representing 16% of all the variants underscoring the impact of SVs in A/M and the importance of including SV interrogation tools such as SVRare^25^ into clinical pipelines. The application of deep learning-based predictors, AlphaMissense and SpliceAI, to large-scale variant datasets provides an effective approach for variant prioritisation. Despite not being explicitly trained on ACMG/AMP criteria, 66.7% (40/60) of the variants flagged by these algorithms, and included in this study, met the pathogenic or likely pathogenic classification. The structural and stability effects of missense variants were further evaluated using Missense3D and molecular dynamic simulations. RMSF analysis revealed local flexibility changes indicative of altered fold stability, as observed for PAX6 variants (Fig.3C, D). While Missense3D predicted no structural damage for *PAX6*:p.Asn139His and *SIX3*, MD simulations indicated destabilisation; conversely, *PORCN*:p.Phe250Ser was predicted as damaging by Missense3D but showed no instability in RMSF, though its previous association with microphthalmia supports pathogenicity^52,55,74^. Such discrepancies highlight that many pathogenic variants act through mechanisms beyond fold stability, such as protein– protein interactions, impaired complex formation, or dysregulated localization, which conventional MD cannot capture^56^. This may also explain the limited correlation between AlphaMissense pathogenicity scores and structural instability predictions, as AlphaMissense integrates broader evolutionary and functional features^75^.

Overall, applying our variant interpretation pipeline could increase the molecular diagnosis rate for the A/M cohort by 12.1% with ACMG guidelines. Despite this effort, 80.4% of the cohort remains unsolved, and compared to other ophthalmic disease cohorts in the 100K GP, the A/M diagnostic yield remains low. Factors contributing to this may include yet unidentified A/M-associated genes, regulatory variants, epigenetic regulation, non-Mendelian inheritance and non-genetic phenocopies, none of which were investigated here.

Altogether, our study advances both the precision and scope of A/M genetic diagnosis by offering a rigorously curated gene panel and a refined analytical framework that integrates structural variant detection and deep learning-based prediction algorithms. In addition, we present additional affected individuals harbouring variants in genes with uncertain associations to A/M, consolidating their affiliation in causing A/M.

## Data Availability

All data produced in the present work are contained in the manuscript.

## Acknowledgements

This research was made possible through access to data in the National Genomic Research Library, which is managed by Genomics England Limited (a company owned by the Department of Health and Social Care). The National Genomic Research Library holds data provided by patients and collected by the NHS as part of their care and data collected as part of their participation in research. The National Genomic Research Library is funded by the National Institute for Health Research and NHS England. The Wellcome Trust Cancer Research UK, and the Medical Research Council have also funded research infrastructure.

This research was supported by the NIHR Moorfields Biomedical Research Centre (GA), the Moorfields Eye Charity Career Development award (GR001155 to RMY), Springboard Awards (GR001210 and GR001662 to RMY), PhD Studentship (GR001661 to MIM), Medical Research Council (MR/X001067/1 to RMY), FONDECYT (1221843 to RMY), Biotechnology and Biological Sciences Research Council (BB/T002212/1 to FF as principal investigator) and the British Heart Foundation grant (RG/F/22/110079 to FF and OGC).

## Competing interests

The authors declare no competing interests.

## Data and resource availability

All relevant data and resources can be found within the article and its supplementary information.

## Author contribution

MM: methodology; formal analysis; investigation; writing – original draft; writing – review and editing; visualisation

LS: formal analysis; investigation; writing

OGC: methodology; formal analysis; investigation; writing SM, LA, RH, AB, MEC, KC, RAS, FF, JY: investigation

GA: methodology; validation; formal analysis; investigation; review and editing; supervision.

RY: conceptualisation; methodology; validation; formal analysis; investigation; resources; writing – original draft; writing – review and editing; visualisation; supervision; project administration; funding acquisition.

---

**Table. 1.** Genes reported to be associated with A/M in literature, and corresponding publications. This panel was generated by aggregating the genes associated by A/M according to the Genomics England PanelApp and the three reviews stated. A 1 within the PanelApp and three review columns, indicates that they have reported that gene as associated with A/M. Every gene above the black threshold has pathogenic variants reported in at least five individuals across two or more families and are deemed to have a confident association with A/M. Genes between the black and red thresholds are deemed to have a weak association with A/M, due to pathogenic variants reported in only one to four individuals, or with family data heavily suggesting an association. Genes with an asterisk (*) appear only in syndromic presentations with sporadic A/M features, indicating these may be incidental and may not represent true associations.

| Gene | [GEL PanelApp] | [Harding and Moosajee, 2019] | [Plaisancie et al., 2019] | [Patel et al., 2019] | Occurance in the<br>four reviews | Number of patients in<br>literature | Number of familes in<br>literature |
| --- | --- | --- | --- | --- | --- | --- | --- |
| SOX2 | 1 | 1 | 1 | 1 | 4 | 161 | 147 |
| PRSS56 | 1 | 1 | 1 | 1 | 4 | 161 | 100 |
| MFRP | 1 | 1 | 1 | 1 | 4 | 129 | 103 |
| OTX2 | 1 | 1 | 1 | 1 | 4 | 104 | 64 |
| BCOR | 1 | 1 | 1 | 1 | 4 | 79 | 66 |
| FOXE3 | 1 | 1 | 1 | 0 | 3 | 75 | 48 |
| VSX2 | 1 | 1 | 1 | 1 | 4 | 74 | 34 |
| TMEM98 | 1 | 1 | 1 | 1 | 4 | 66 | 13 |
| RAB3GAP1 | 1 | 1 | 1 | 1 | 4 | 59 | 47 |
| ALDH1A3 | 1 | 1 | 1 | 1 | 4 | 59 | 39 |
| STRA6 | 1 | 1 | 1 | 1 | 4 | 48 | 32 |
| PORCN | 0 | 1 | 1 | 1 | 3 | 38 | 32 |
| PAX6 | 1 | 1 | 1 | 1 | 4 | 35 | 28 |
| MYRF | 1 | 0 | 0 | 0 | 1 | 35 | 14 |
| GJA1 | 0 | 1 | 0 | 1 | 2 | 30 | 17 |
| HCCS | 1 | 1 | 1 | 1 | 4 | 29 | 27 |
| BMP4 | 1 | 1 | 1 | 1 | 4 | 27 | 26 |
| SIX6 | 1 | 1 | 1 | 1 | 4 | 27 | 20 |
| SMCHD1 | 0 | 1 | 1 | 0 | 2 | 26 | 25 |
| RAB3GAP2 | 1 | 1 | 1 | 1 | 4 | 25 | 18 |
| GJA8 | 0 | 0 | 1 | 0 | 1 | 25 | 20 |
| MAB21L2 | 1 | 1 | 1 | 1 | 4 | 23 | 20 |
| CHD7 | 0 | 1 | 1 | 0 | 2 | 23 | 23 |
| RARB | 1 | 1 | 1 | 1 | 4 | 22 | 17 |
| TFAP2A | 0 | 1 | 1 | 1 | 3 | 19 | 17 |
| RAX | 1 | 1 | 1 | 1 | 4 | 18 | 17 |
| PXDN | 0 | 1 | 1 | 0 | 2 | 17 | 12 |
| SMOC1 | 1 | 1 | 1 | 1 | 4 | 15 | 10 |
| GDF6 | 1 | 1 | 1 | 1 | 3 | 15 | 15 |
| C12ORF57 | 0 | 1 | 1 | 1 | 3 | 14 | 13 |
| NHS | 0 | 1 | 0 | 0 | 1 | 14 | 14 |
| YAP1 | 0 | 1 | 1 | 0 | 2 | 13 | 6 |
| TBC1D20 | 1 | 0 | 1 | 1 | 3 | 12 | 7 |
| RAB18 | 1 | 1 | 1 | 0 | 3 | 12 | 7 |
| CRYAA | 0 | 1 | 0 | 1 | 2 | 11 | 6 |
| EPHA2 | 0 | 0 | 0 | 0 | 0 | 11 | 4 |
| PAX2 | 1 | 1 | 1 | 0 | 3 | 9 | 9 |
| GDF3 | 1 | 1 | 1 | 1 | 3 | 9 | 8 |
| ERCC6 | 1 | 1 | 0 | 0 | 1 | 9 | 8 |
| PRR12 | 0 | 0 | 0 | 0 | 0 | 9 | 8 |
| TMX3 | 0 | 1 | 1 | 1 | 3 | 8 | 8 |
| NAA10 | 0 | 1 | 1 | 1 | 3 | 8 | 3 |
| COL4A1 | 1 | 1 | 1 | 0 | 3 | 8 | 7 |
| LRP5 | 0 | 1 | 0 | 0 | 1 | 8 | 6 |
| CRPPA | 0 | 1 | 0 | 0 | 1 | 8 | 8 |
| PTCH1 | 0 | 1 | 1 | 1 | 3 | 7 | 7 |
| KMT2D | 1 | 1 | 0 | 0 | 2 | 7 | 7 |
| PITX3 | 1 | 1 | 1 | 0 | 2 | 7 | 6 |
| ZEB2 | 0 | 1 | 0 | 0 | 1 | 7 | 7 |
| FAT1 | 1 | 0 | 0 | 0 | 1 | 7 | 5 |
| TENM3 | 1 | 1 | 1 | 1 | 4 | 6 | 5 |
| ATOH7 | 0 | 1 | 1 | 1 | 3 | 6 | 2 |
| SHH | 1 | 1 | 0 | 1 | 3 | 6 | 6 |
| FAM111A | 0 | 1 | 0 | 1 | 2 | 6 | 5 |
| SLC24A5 | 0 | 1 | 0 | 0 | 1 | 6 | 6 |
| FKTN | 0 | 1 | 0 | 0 | 1 | 6 | 6 |
| ALX1 | 0 | 1 | 0 | 0 | 1 | 6 | 3 |
| ABCB6 | 1 | 1 | 1 | 1 | 3 | 5 | 5 |
| TBC1D32 | 0 | 1 | 0 | 1 | 2 | 5 | 5 |
| CRYBA4 | 0 | 1 | 0 | 1 | 2 | 5 | 4 |
| HMX1 | 0 | 1 | 0 | 1 | 2 | 5 | 2 |
| MITF | 0 | 1 | 1 | 0 | 2 | 5 | 5 |
| POMT1 | 0 | 1 | 0 | 0 | 1 | 5 | 5 |
| NDP | 0 | 1 | 0 | 0 | 1 | 5 | 4 |
| MAPRE2 | 0 | 1 | 0 | 0 | 1 | 5 | 5 |
| DOCK6 | 0 | 1 | 0 | 0 | 1 | 5 | 5 |
| ERCC2 | 1 | 0 | 0 | 0 | 0 | 5 | 5 |
| TUBGCP4 | 1 | 0 | 0 | 0 | 0 | 5 | 4 |
| KERA | 0 | 0 | 0 | 0 | 0 | 5 | 2 |
| FREM1 | 1 | 1 | 1 | 1 | 4 | 4 | 4 |
| RBP4 | 0 | 1 | 1 | 1 | 3 | 4 | 4 |
| FRAS1 | 1 | 1 | 1 | 0 | 3 | 4 | 3 |
| SALL4 | 0 | 1 | 1 | 1 | 3 | 4 | 3 |
| FREM2 | 1 | 1 | 0 | 0 | 2 | 4 | 4 |
| PQBP1 | 0 | 1 | 0 | 1 | 2 | 4 | 2 |
| FOXC1 | 1 | 1 | 0 | 0 | 2 | 4 | 4 |
| TUBB | 0 | 1 | 0 | 0 | 1 | 4 | 3 |
| SRD5A3 | 0 | 1 | 0 | 0 | 1 | 4 | 4 |
| CAPN15 | 1 | 0 | 0 | 0 | 1 | 4 | 3 |
| PLK4 | 1 | 0 | 0 | 0 | 0 | 4 | 2 |
| B3GLCT | 1 | 0 | 0 | 0 | 0 | 4 | 4 |
| Six3 | 0 | 1 | 1 | 1 | 3 | 3 | 3 |
| BMP7 | 0 | 1 | 1 | 1 | 3 | 3 | 3 |
| B3GALNT2 | 0 | 1 | 0 | 0 | 1 | 3 | 3 |
| SALL1 | 0 | 1 | 0 | 0 | 1 | 3 | 2 |
| RPGRIP1L | 0 | 1 | 0 | 0 | 1 | 3 | 2 |
| POMGNT1 | 0 | 1 | 0 | 0 | 1 | 3 | 3 |
| KIF11 | 0 | 1 | 0 | 0 | 1 | 3 | 3 |
| CRYBB2 | 0 | 0 | 0 | 1 | 1 | 3 | 3 |
| FKRP* | 0 | 1 | 0 | 0 | 1 | 3 | 3 |
| POMT2 | 0 | 1 | 0 | 0 | 1 | 3 | 3 |
| C16orf62 | 1 | 0 | 0 | 0 | 0 | 3 | 3 |
| SMO | 1 | 1 | 1 | 0 | 3 | 2 | 2 |
| ERCC5* | 1 | 1 | 0 | 1 | 2 | 2 | 2 |
| FOXL2* | 0 | 1 | 0 | 1 | 2 | 2 | 2 |
| PDE6D | 0 | 1 | 0 | 1 | 2 | 2 | 1 |
| GLI2* | 0 | 1 | 0 | 0 | 1 | 2 | 2 |
| CEP290* | 0 | 1 | 0 | 0 | 1 | 2 | 2 |
| RERE* | 0 | 1 | 0 | 0 | 1 | 2 | 2 |
| OLFM2 | 0 | 0 | 1 | 0 | 1 | 2 | 2 |
| MAF* | 0 | 0 | 1 | 0 | 1 | 2 | 2 |
| *DPYD\** | 0 | 0 | 0 | 1 | 1 | 2 | 2 |
| *SMG9* | 0 | 1 | 0 | 0 | 1 | 2 | 2 |
| *ACTB\** | 0 | 0 | 1 | 0 | 1 | 2 | 2 |
| *SNX3* | 0 | 1 | 0 | 0 | 1 | 2 | 2 |
| *FANCL\** | 0 | 1 | 0 | 0 | 1 | 2 | 2 |
| *NR6A1* | 0 | 0 | 0 | 0 | 0 | 2 | 2 |
| *CYP1B1\** | 1 | 0 | 0 | 0 | 0 | 2 | 2 |
| *FNBP4* | 0 | 1 | 1 | 1 | 3 | 1 | 1 |
| *VAX1* | 1 | 1 | 1 | 1 | 3 | 1 | 1 |
| *ERCC1* | 1 | 1 | 0 | 1 | 2 | 1 | 1 |
| *PITX2\** | 1 | 0 | 0 | 0 | 1 | 1 | 1 |
| *IPO13* | 0 | 1 | 0 | 0 | 1 | 1 | 1 |
| *SCLT1\** | 0 | 0 | 0 | 1 | 1 | 1 | 1 |
| *TCTN2\** | 0 | 1 | 0 | 0 | 1 | 1 | 1 |
| *CSPP1\** | 0 | 1 | 0 | 0 | 1 | 1 | 1 |
| *TMEM216\** | 0 | 1 | 0 | 0 | 1 | 1 | 1 |
| *ACTG1\** | 0 | 0 | 1 | 0 | 1 | 1 | 1 |
| *GTF2H5\** | 1 | 0 | 0 | 0 | 0 | 1 | 1 |
| *BMPR1B\** | 0 | 0 | 0 | 0 | 0 | 1 | 1 |
| *HMGB3* | 0 | 1 | 1 | 1 | 3 | 0 | 0 |
| *HDAC6* | 1 | 1 | 0 | 0 | 1 | 0 | 0 |
| *LRP2\** | 0 | 0 | 1 | 0 | 1 | 0 | 0 |
| *DAG1\** | 0 | 1 | 0 | 0 | 1 | 0 | 0 |
| *COX7B\** | 0 | 0 | 1 | 0 | 1 | 0 | 0 |
| *GRIP1\** | 1 | 0 | 0 | 0 | 1 | 0 | 0 |
| *PAX3\** | 0 | 0 | 1 | 0 | 1 | 0 | 0 |
| *SOX10\** | 0 | 0 | 1 | 0 | 1 | 0 | 0 |

**Table. 2.**
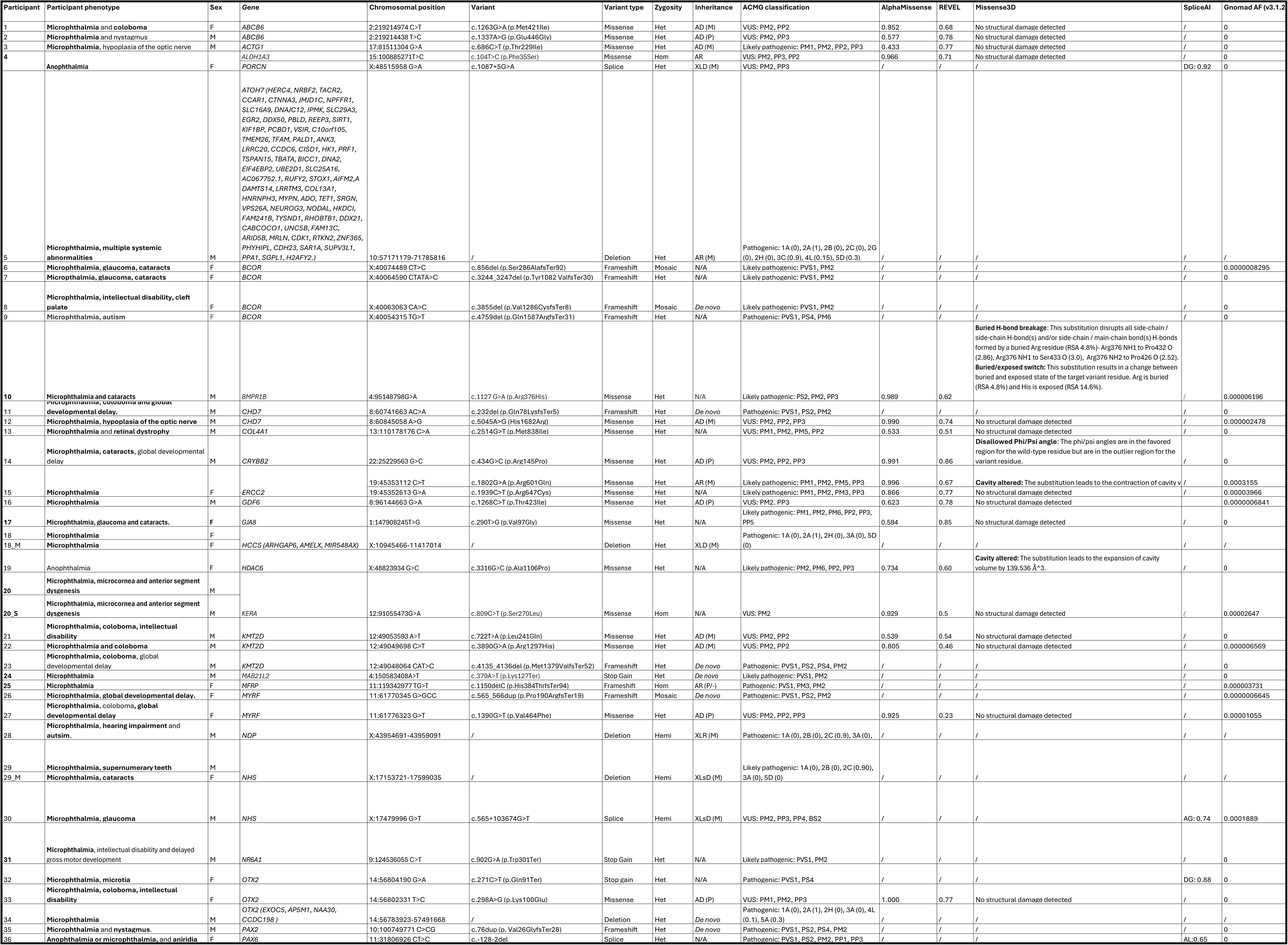
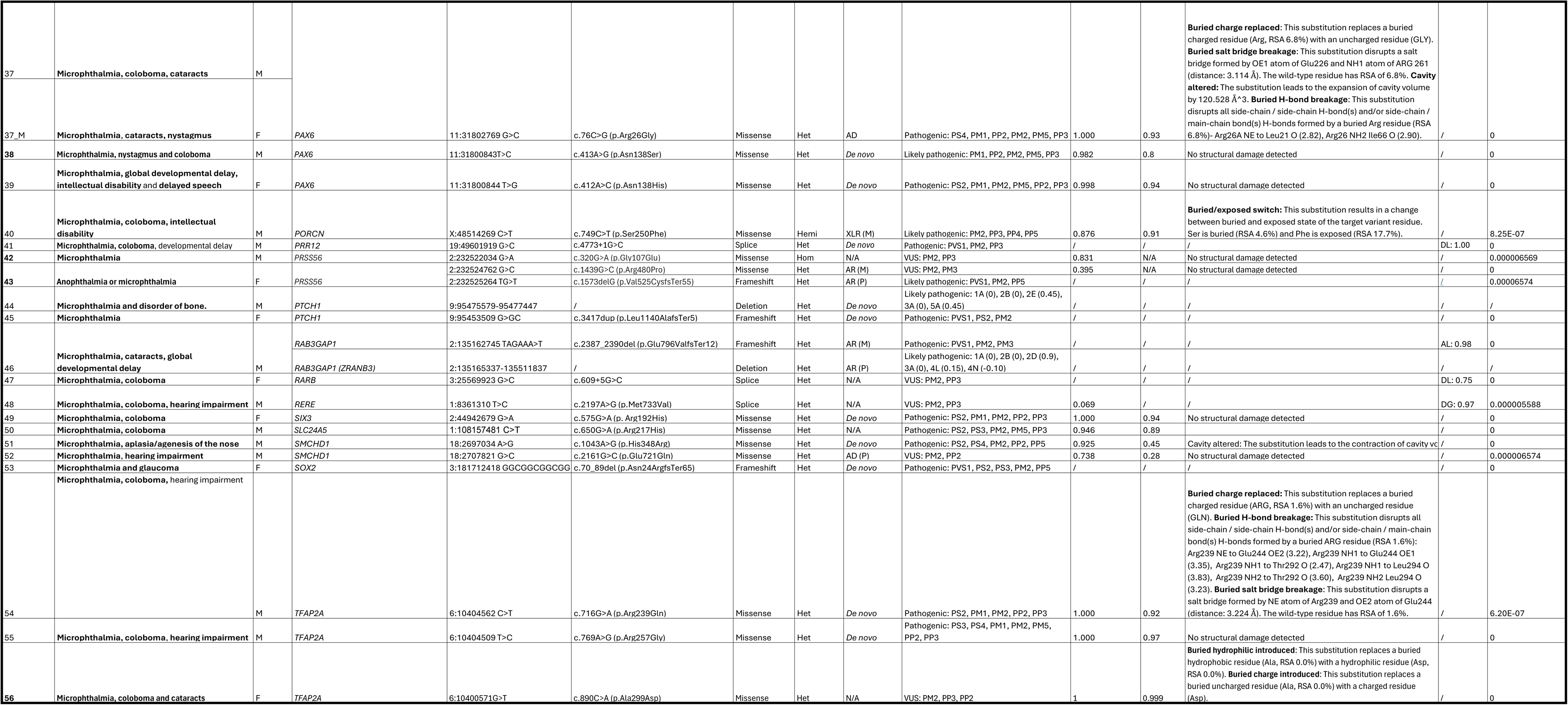
Candidate variants identified within the 100KGP anophthalmia and microphthalmia cohort. Participant numbers in bold are ones who have been previously moleculary diagnosed. Phenotypes in bold are ones which are associated with the respective gene. Inheritance of the variant is stated whether it was inherited maternally (M), or paternally (P), de novo, or whether there is insufficient parental data (N/A). AD: Autosomal dominant, AF: Allele frequency, AG: Acceptor gain, AL: Acceptor loss, AR: Autosomal recessive, DG: Donor gain, DL: Donor loss, Het: Heterozygous, Hom: Homozygous, VUS: variant of uncertain significance, XLD: X-linked dominant, XLR: X-linked recessive, XLsD: X-linked semi-dominant

**Table. 3.**
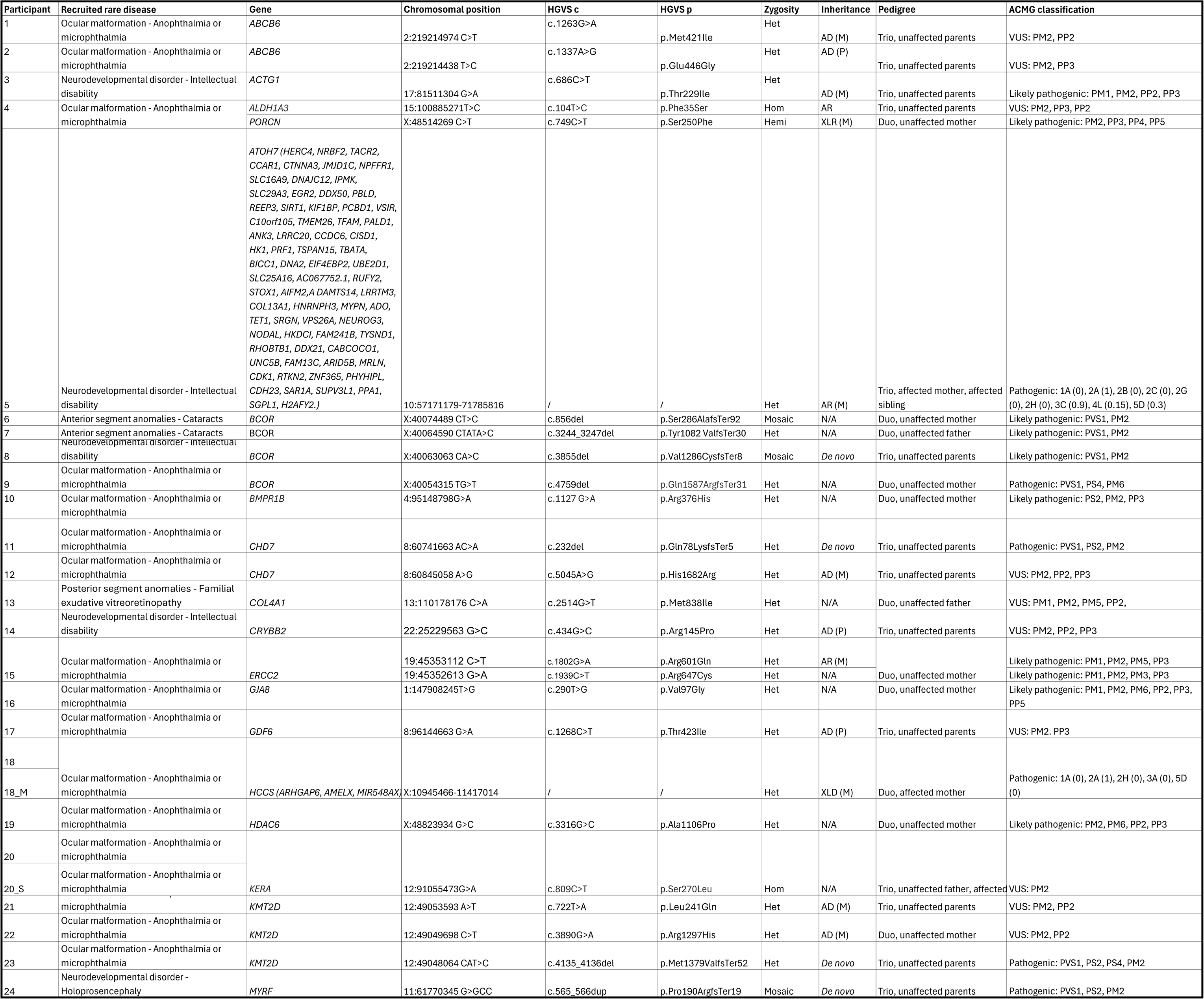
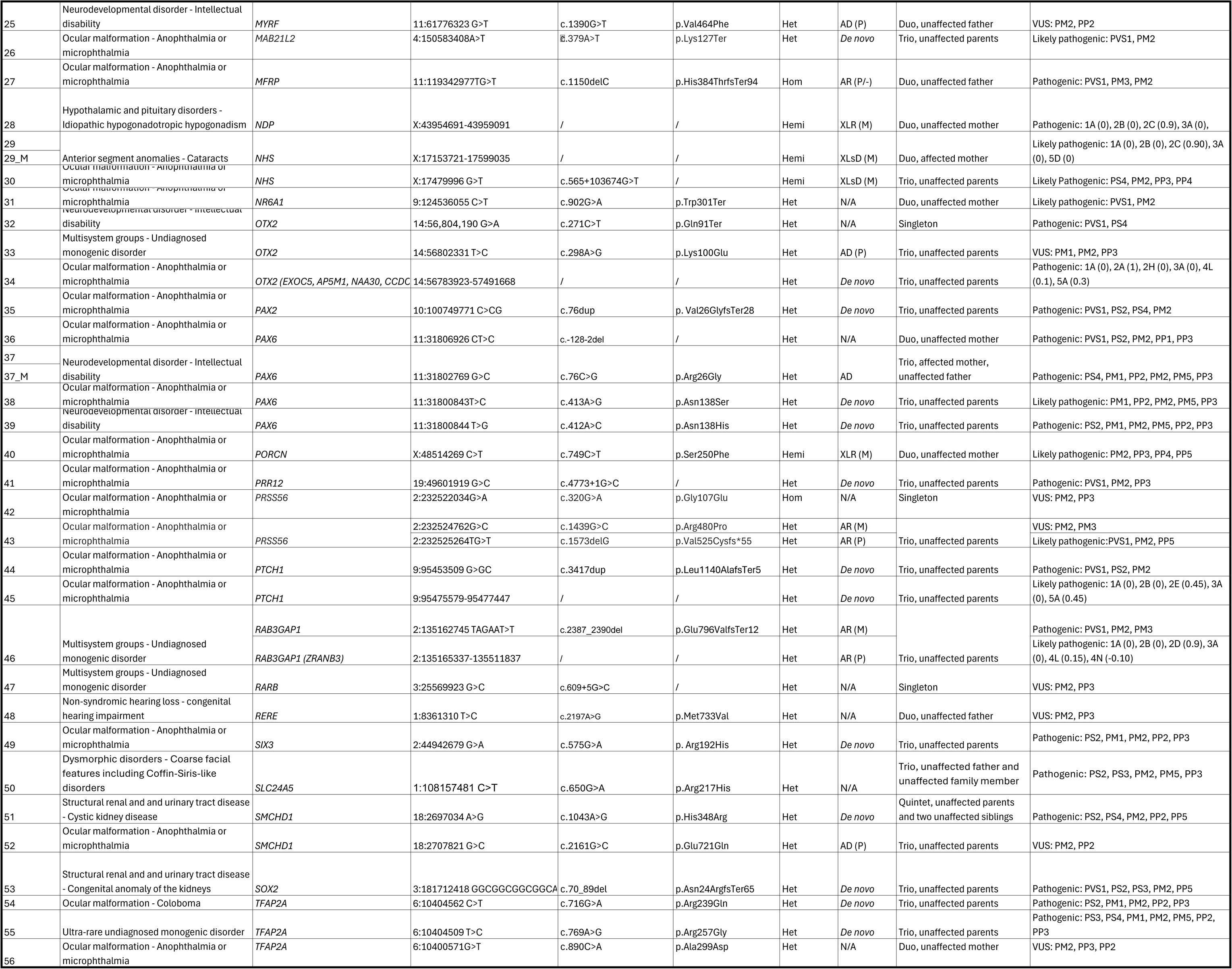
Recruitment and pedigree information of participants in the 100KGP anophthalmia and microphthalmia cohort. Phenotypes in bold are ones which are associated with the respective gene. Recruited disease refers to the disorder for which the family was recruited to the 100K GP. Inheritance of the variant is stated whether it was inherited maternally (M), or paternally (P), de novo, or whether there is insufficient parental data (N/A). AD: Autosomal dominant, AF: Allele frequency, AG: Acceptor gain, AL: Acceptor loss, AR: Autosomal recessive, DG: Donor gain, DL: Donor loss, Het: Heterozygous, Hom: Homozygous, VUS: variant of uncertain significance, XLD: X-linked dominant, XLR: X-linked recessive, XLsD: X-linked semi-dominant.

## Notes

### Competing Interest Statement

The authors have declared no competing interest.

### Author Declarations

The East of England, Cambridge South Research Ethics Committee of the UK Health Research Authority gave ethical approval for this work (REC reference: 14/EE/1112). Access to the data was granted under Genomics England's Data Access Agreement (Project ID: RR439).

### Summary of Updates

The author affiliations have been updated.

## References

1. Smedley D, Smith KR, Martin A, et al. 100,000 Genomes Pilot on Rare-Disease Diagnosis in Health Care — Preliminary Report. N Engl J Med. 2021;385(20):1868–1880. doi:10.1056/nejmoa2035790

2. Shieh JTC. Genomic technologies to improve variation identification in undiagnosed diseases. Pediatr Neonatol. 2023;64:S18–S21. doi:10.1016/j.pedneo.2022.10.002

3. Turro E, Astle WJ, Megy K, et al. Whole-genome sequencing of patients with rare diseases in a national health system. Nature. 2020;583(7814):96–102. doi:10.1038/s41586-020-2434-2

4. Marwaha S, Knowles JW, Ashley EA. A guide for the diagnosis of rare and undiagnosed disease: beyond the exome. Genome Med. 2022;14(1). doi:10.1186/s13073-022-01026-w

5. Schneider A, Bardakjian T, Reis LM, Tyler RC, Semina EV. Novel SOX2 mutations and genotype-phenotype correlation in anophthalmia and microphthalmia. Am J Med Genet A. 2009;149A(12):2706–2715. doi:10.1002/ajmg.a.33098

6. Shah SP, Taylor AE, Sowden JC, et al. Anophthalmos, Microphthalmos, and Coloboma in the United Kingdom: Clinical Features, Results of Investigations, and Early Management. Ophthalmology. 2012;119(2):362–368. doi:10.1016/j.ophtha.2011.07.039

7. Chassaing N, Causse A, Vigouroux A, et al. Molecular findings and clinical data in a cohort of 150 patients with anophthalmia/microphthalmia. Clin Genet. 2013;86(4):326–334. doi:10.1111/cge.12275

8. Gerth-Kahlert C, Williamson KA, Ansari M, et al. Clinical and mutation analysis of 51 probands with anophthalmia and/or severe microphthalmia from a single center. Mol Genet Genomic Med. 2013;1(1):15–31. doi:10.1002/mgg3.2

9. Slavotinek AM, Garcia ST, Chandratillake G, et al. Exome sequencing in 32 patients with anophthalmia/microphthalmia and developmental eye defects. Clin Genet. 2015;88(5):468–473. doi:10.1111/cge.12543

10. Patel A, Hayward JD, Tailor V, et al. The Oculome Panel Test. Ophthalmology. 2019;126(6):888–907. doi:10.1016/j.ophtha.2018.12.050

11. Harding P, Moosajee M. The Molecular Basis of Human Anophthalmia and Microphthalmia. J Dev Biol. 2019;7(3). doi:10.3390/jdb7030016

12. Plaisancié J, Ceroni F, Holt R, et al. Genetics of anophthalmia and microphthalmia. Part 1: Non-syndromic anophthalmia/microphthalmia. Hum Genet. 2019;138(8-9):799–830. doi:10.1007/s00439-019-01977-y

13. Harding P, Gore S, Malka S, Rajkumar J, Oluonye N, Moosajee M. Real-world clinical and molecular management of 50 prospective patients with microphthalmia, anophthalmia and/or ocular coloboma. Br J Ophthalmol. 2022;107(12):bjo-2022. doi:10.1136/bjo-2022-321991

14. Reis LM, Semina EV. Conserved genetic pathways associated with microphthalmia, anophthalmia, and coloboma. Birth Defects Res Part C Embryo Today Rev. 2015;105(2):96–113. doi:10.1002/bdrc.21097

15. Richardson R, Sowden J, Gerth-Kahlert C, Moore AT, Mariya Moosajee. Clinical utility gene card for: Non-Syndromic Microphthalmia Including Next-Generation Sequencing-Based Approaches. Eur J Hum Genet. 2017;25(4):512–512. doi:10.1038/ejhg.2016.201

16. Slavotinek A. Genetics of anophthalmia and microphthalmia. Part 2: Syndromes associated with anophthalmia–microphthalmia. Hum Genet. 2018;138(8-9):831–846. doi:10.1007/s00439-018-1949-1

17. Goyal S, Tibrewal S, Ratna R, Vanita V. Genetic and environmental factors contributing to anophthalmia and microphthalmia: Current understanding and future directions. World J Clin Pediatr. 2025;14(2). doi:10.5409/wjcp.v14.i2.101982

18. The National Genomic Research Library v5.1, Genomics England. 2020. doi:10.6084/m9.figshare.4530893/7

19. Anophthalmia or microphthalmia (Version 1.51). Genomicsengland.co.uk. 2016. https://panelapp.genomicsengland.co.uk/panels/34/

20. Harding P, Toms M, Schiff E, et al. EPHA2 Segregates with Microphthalmia and Congenital Cataracts in Two Unrelated Families. Int J Mol Sci. 2021;22(4):2190. doi:10.3390/ijms22042190

21. Courdier C, Gemahling A, Guindolet D, et al. EPHA2 biallelic disruption causes syndromic complex microphthalmia with iris hypoplasia. Eur J Med Genet. 2022;65(10):104574. doi:10.1016/j.ejmg.2022.104574

22. Neelathi UM, Ullah E, George A, et al. Variants in NR6A1 cause a novel oculo-vertebral-renal (OVR) syndrome. PubMed. Published online 2024. doi:10.1101/2024.11.09.24316578

23. Reis LM, Costakos D, Wheeler PG, et al. Dominant variants in PRR12 result in unilateral or bilateral complex microphthalmia. Clin Genet. 2021;99(3):437–442. doi:10.1111/cge.13897

24. Chowdhury F, Wang L, Al-Raqad M, et al. Haploinsufficiency of PRR12 causes a spectrum of neurodevelopmental, eye, and multisystem abnormalities. Genet Med. 2021;23(7):1234–1245. doi:10.1038/s41436-021-01129-6

25. Yu J, Szabo A, Pagnamenta AT, et al. SVRare: discovering disease-causing structural variants in the 100K Genomes Project. MedRxiv Cold Spring Harb Lab. Published online 2021. doi:10.1101/2021.10.15.21265069

26. Karczewski KJ, Francioli LC, Tiao G, et al. The mutational constraint spectrum quantified from variation in 141,456 humans. Nature. 2020;581(7809):434–443. doi:10.1038/s41586-020-2308-7

27. Robinson JT, Thorvaldsdóttir H, Winckler W, et al. Integrative genomics viewer. Nat Biotechnol. 2011;29(1):24–26. doi:10.1038/nbt.1754

28. Ioannidis NM, Rothstein JH, Pejaver V, et al. REVEL: An Ensemble Method for Predicting the Pathogenicity of Rare Missense Variants. Am J Hum Genet. 2016;99(4):877–885. doi:10.1016/j.ajhg.2016.08.016

29. Ittisoponpisan S, Islam SA, Khanna T, Alhuzimi E, David A, Sternberg MJE. Can Predicted Protein 3D Structures Provide Reliable Insights into whether Missense Variants Are Disease Associated? J Mol Biol. 2019;431(11):2197–2212. doi:10.1016/j.jmb.2019.04.009

30. Zenteno JC, Perez-Cano HJ, Aguinaga M. Anophthalmia-esophageal atresia syndrome caused by an SOX2 gene deletion in monozygotic twin brothers with markedly discordant phenotypes. Am J Med Genet A. 2006;140A(18):1899–1903. doi:10.1002/ajmg.a.31384

31. Schneider A, Bardakjian TM, Zhou J, et al. Familial recurrence ofSOX2anophthalmia syndrome: Phenotypically normal mother with two affected daughters. Am J Med Genet A. 2008;146A(21):2794–2798. doi:10.1002/ajmg.a.32384

32. Suzuki J, Azuma N, Dateki S, et al. Mutation spectrum and phenotypic variation in nine patients with SOX2 abnormalities. J Hum Genet. 2014;59(6):353–356. doi:10.1038/jhg.2014.34

33. Cheon CK, Ko JM. Kabuki syndrome: clinical and molecular characteristics. Korean J Pediatr. 2015;58(9):317. doi:10.3345/kjp.2015.58.9.317

34. Bögershausen N, Gatinois V, Riehmer V, et al. Mutation Update for Kabuki Syndrome GenesKMT2DandKDM6Aand Further Delineation of X-Linked Kabuki Syndrome Subtype 2. Hum Mutat. 2016;37(9):847–864. doi:10.1002/humu.23026

35. Errichiello E, Gorgone C, Giuliano L, et al. SOX2: Not always eye malformations. Severe genital but no major ocular anomalies in a female patient with the recurrent c.70del20 variant. Eur J Med Genet. 2018;61(6):335–340. doi:10.1016/j.ejmg.2018.01.011

36. Zhang Y, Zhang X, Long R, Yu L. A novel deletion mutation of the SOX2 gene in a child of Chinese origin with congenital bilateral anophthalmia and sensorineural hearing loss. J Genet. 2018;97(4):1007–1011. doi:10.1007/s12041-018-0970-4

37. Xiong HY, Shi YQ, Zhong C, et al. Detection of De Novo PAX2 Variants and Phenotypes in Chinese Population: A Single-Center Study. Front Genet. 2022;13. doi:10.3389/fgene.2022.799562

38. Sanyanusin P, McNoe LA, Sullivan MJ, Weaver RG, Eccles MR. Mutation of PAX2 in two siblings with renal-coloboma syndrome. Hum Mol Genet. 1995;4(11):2183–2184. doi:10.1093/hmg/4.11.2183

39. Ford B, Rupps R, Lirenman D, et al. Renal-coloboma syndrome: Prenatal detection and clinical spectrum in a large family. Am J Med Genet. 2001;99(2). doi:10.1002/1096-8628(2000)9999:999%3C00::AID-AJMG1143%3E3.0.CO;2-F

40. Garnai SJ, Brinkmeier ML, Emery B, et al. Variants in myelin regulatory factor (MYRF) cause autosomal dominant and syndromic nanophthalmos in humans and retinal degeneration in mice. Anderson MG, ed. PLOS Genet. 2019;15(5):e1008130. doi:10.1371/journal.pgen.1008130

41. Rossetti LZ, Glinton K, Yuan B, et al. Review of the phenotypic spectrum associated with haploinsufficiency of MYRF. Am J Med Genet A. 2019;179(7):1376–1382. doi:10.1002/ajmg.a.61182

42. Siggs OM, Awadalla MS, Souzeau E, et al. The genetic and clinical landscape of nanophthalmos and posterior microphthalmos in an Australian cohort. Clin Genet. 2020;97(5):764–769. doi:10.1111/cge.13722

43. Hilton E, Johnston JJ, Whalen S, et al. BCOR analysis in patients with OFCD and Lenz microphthalmia syndromes, mental retardation with ocular anomalies, and cardiac laterality defects. Eur J Hum Genet. 2009;17(10):1325–1335. doi:10.1038/ejhg.2009.52

44. Ragge N, Isidor B, Bitoun P, et al. Expanding the phenotype of the X-linked BCOR microphthalmia syndromes. Hum Genet. 2018;138(8-9):1051–1069. doi:10.1007/s00439-018-1896-x

45. Riggs ER, Andersen EF, Cherry AM, et al. Technical standards for the interpretation and reporting of constitutional copy-number variants: a joint consensus recommendation of the American College of Medical Genetics and Genomics (ACMG) and the Clinical Genome Resource (ClinGen). Genet Med. 2019;22. doi:10.1038/s41436-019-0686-8

46. Yu J, Liao PJ, Xu W, et al. Structural model of human PORCN illuminates disease-associated variants and drug-binding sites. J Cell Sci. 2021;134(24):jcs259383. doi:10.1242/jcs.259383

47. Asai-Coakwell M, French CR, Ye M, et al. Incomplete penetrance and phenotypic variability characterize Gdf6-attributable oculo-skeletal phenotypes. Hum Mol Genet. 2009;18(6):1110–1121. doi:10.1093/hmg/ddp008

48. Cuvertino S, Hartill V, Colyer A, et al. A restricted spectrum of missense KMT2D variants cause a multiple malformations disorder distinct from Kabuki syndrome. Genet Med. 2020;22(5):867–877. doi:10.1038/s41436-019-0743-3

49. Gregory LC, Gergics P, Nakaguma M, et al. The phenotypic spectrum associated with OTX2 mutations in humans. Eur J Endocrinol. 2021;185(1):121–135. doi:10.1530/EJE-20-1453

50. Shaw ND, Brand H, Kupchinsky ZA, et al. SMCHD1 mutations associated with a rare muscular dystrophy can also cause isolated arhinia and Bosma arhinia microphthalmia syndrome. 2017;49(2):238–248. doi:10.1038/ng.3743

51. Zentner GE, Layman WS, Martin DM, Scacheri PC. Molecular and phenotypic aspects of CHD7 mutation in CHARGE syndrome. Am J Med Genet A. 2010;152A(3):674–686. doi:10.1002/ajmg.a.33323

52. Madan S, Liu W, Lu JT, et al. A non-mosaic PORCN mutation in a male with severe congenital anomalies overlapping focal dermal hypoplasia. Mol Genet Metab Rep. 2017;12:57–61. doi:10.1016/j.ymgmr.2017.06.002

53. Wawrocka A, Walczak-Sztulpa J, Pawlak M, Gotz-Wieckowska A, Krawczynski MR. Non-syndromic anophthalmia/microphthalmia can be caused by a PORCN variant inherited in X-linked recessive manner. Am J Med Genet - Part A. 2020;185(1):250–255. doi:10.1002/ajmg.a.61938

54. Kesim Y, Ceroni F, Damián A, et al. Clinical and genetic analysis further delineates the phenotypic spectrum of ALDH1A3-related anophthalmia and microphthalmia. Eur J Hum Genet. 2023;31(10):1175–1180. doi:10.1038/s41431-023-01342-8

55. Buric F, Viknander S, Fu X, et al. Amino acid sequence encodes protein abundance shaped by protein stability at reduced synthesis cost. Protein Sci. 2024;34(1). doi:10.1002/pro.5239

56. Beltran A, Jiang X, Shen Y, Lehner B. Site-saturation mutagenesis of 500 human protein domains. Nature. 2025;637. doi:10.1038/s41586-024-08370-4

57. Ragge NK, Brown AG, Poloschek CM, et al. Heterozygous Mutations of OTX2 Cause Severe Ocular Malformations. Am J Hum Genet. 2005;76(6):1008–1022. doi:10.1086/430721

58. Chassaing N, Sorrentino S, Davis EE, et al. OTX2mutations contribute to the otocephaly-dysgnathia complex. J Med Genet. 2012;49(6):373–379. doi:10.1136/jmedgenet-2012-100892

59. Williamson KA, FitzPatrick DR. The genetic architecture of microphthalmia, anophthalmia and coloboma. Eur J Med Genet. 2014;57(8):369–380. doi:10.1016/j.ejmg.2014.05.002

60. Li X, Xiao H, Su Y, et al. Clinical features of patients with mutations in genes for nanophthalmos. Br J Ophthalmol. 2024;108(12):1679–1687. doi:10.1136/bjo-2023-324931

61. Kondo D, Noguchi A, Tamura H, et al. Elevated Urinary Levels of 8-Hydroxy-2′-deoxyguanosine in a Japanese Child of Xeroderma Pigmentosum/Cockayne Syndrome Complex with Infantile Onset of Nephrotic Syndrome. Tohoku J Exp Med. 2016;239(3):231–235. doi:10.1620/tjem.239.231

62. Graham JM, Anyane-Yeboa K, Raams A, et al. Cerebro-Oculo-Facio-Skeletal Syndrome with a Nucleotide Excision–Repair Defect and a Mutated XPD Gene, with Prenatal Diagnosis in a Triplet Pregnancy. Am J Hum Genet. 2001;69(2):291–300. doi:10.1086/321295

63. Farmaki E, Nedelkopoulou N, Delli F, Sarafidis K, Zafeiriou DI. Brittle Hair, Photosensitivity, Brain Hypomyelination and Immunodeficiency: Clues to Trichothiodystrophy. Indian J Pediatr. 2017;84(1):89–90. doi:10.1007/s12098-016-2209-9

64. Janani R, Chermakani P, Sundaresan P, Shetty S, Rai K. A rare case in a child with mild trichothiodystrophy associated with ERCC2 gene. Indian J Ophthalmol - Case Rep. 2022;2(4):962–964. doi:10.4103/ijo.IJO_1217_22

65. Horibata K, Kono S, Ishigami C, et al. Constructive rescue of TFIIH instability by an alternative isoform of XPD derived from a mutated XPD allele in mild but not severe XP-D/CS. J Hum Genet. 2015;60(5):259–265. doi:10.1038/jhg.2015.18

66. Berger W, Meindl A, Van De Pol TJR, et al. Isolation of a candidate gene for Norrie disease by positional cloning. Nat Genet. 1992;1(3):199–203. doi:10.1038/ng0692-199

67. Payabvash S, Anderson JS, Nascene DR. Bilateral persistent fetal vasculature due to a mutation in the Norrie disease protein gene. Neuroradiol J. 2015;28(6):623–627. doi:10.1177/1971400915609350

68. Deml B, Reis LM, Lemyre E, Clark RD, Kariminejad A, Semina EV. Novel mutations in PAX6, OTX2 and NDP in anophthalmia, microphthalmia and coloboma. Eur J Hum Genet. 2015;24(4):535–541. doi:10.1038/ejhg.2015.155

69. Lin M, Lu Y, Sui Y, et al. A novel c.287G>T *NDP* missense mutation in a Chinese family with Norrie disease. Ophthalmic Genet. 2020;41(4):338–340. doi:10.1080/13816810.2020.1759106

70. Khalifa O, Al-Sahlawi Z, Imtiaz F, et al. Variable expression pattern in Donnai-Barrow syndrome: Report of two novel LRP2 mutations and review of the literature. Eur J Med Genet. 2015;58(5):293–299. doi:10.1016/j.ejmg.2014.12.008

71. Chassaing N, Siani V, Carles D, et al. X-linked dominant chondrodysplasia with platyspondyly, distinctive brachydactyly, hydrocephaly, and microphthalmia. Am J Med Genet A. 2005;136A(4):307–312. doi:10.1002/ajmg.a.30570

72. Simon D, Laloo B, Barillot M, et al. A mutation in the 3′-UTR of the *HDAC6* gene abolishing the post-transcriptional regulation mediated by hsa-miR-433 is linked to a new form of dominant X-linked chondrodysplasia. Hum Mol Genet. 2010;19(10):2015–2027. doi:10.1093/hmg/ddq083

73. Kantarci S, Casavant D, Prada C, et al. Findings from aCGH in patients with congenital diaphragmatic hernia (CDH): A possible locus for Fryns syndrome. Am J Med Genet A. 2006;140A(1):17–23. doi:10.1002/ajmg.a.31025

74. Frisk S, Grandpeix-Guyodo C, Silwerfeldt KP, et al. Goltz syndrome in males: A clinical report of a male patient carrying a novel PORCN variant and a review of the literature. Clin Case Rep. 2018;6(11):2103–2110. doi:10.1002/ccr3.1783

75. Cheng J, Novati G, Pan J, et al. Accurate proteome-wide missense variant effect prediction with AlphaMissense. Science. 2023;381(6664). doi:10.1126/science.adg7492

