## Supplemental Figures for "Refining the genetic landscape of anophthalmia and microphthalmia: a comprehensive framework with deep learning and updated gene panels"

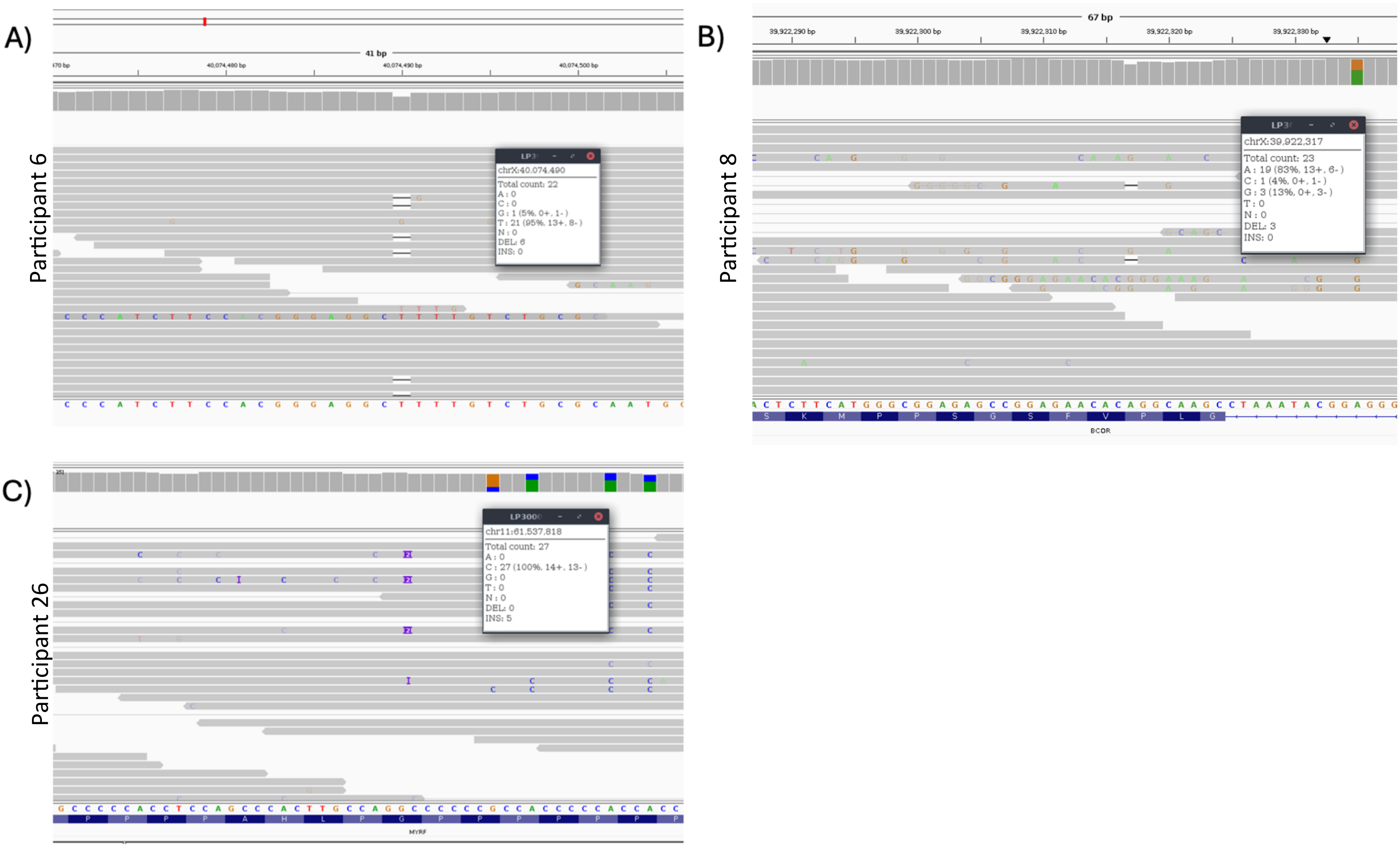


**Supplementary Figure 1.** Mosaic variant expression in A/M participant

A) Participant 6 expresses a mosaic inheritance of the *BCOR* p.Ser286AlafsTer92 variant, occurring in 6 of the reads out of 28 (21.4%). B) Participant 8 expresses a mosaic inheritance of the *BCOR* p.Val1286CysfsTer8 variant, occurring in 3 of the reads out of 26 (11.5%). Note that the chromosomal position is given in hg17. C) Participant 26 expresses a mosaic inheritance of the *MYRF* Pro190ArgfsTer19 variant, occurring in 5 of the reads out of 27 (18.5%). Note that the chromosomal position is given in hg17 assembly.

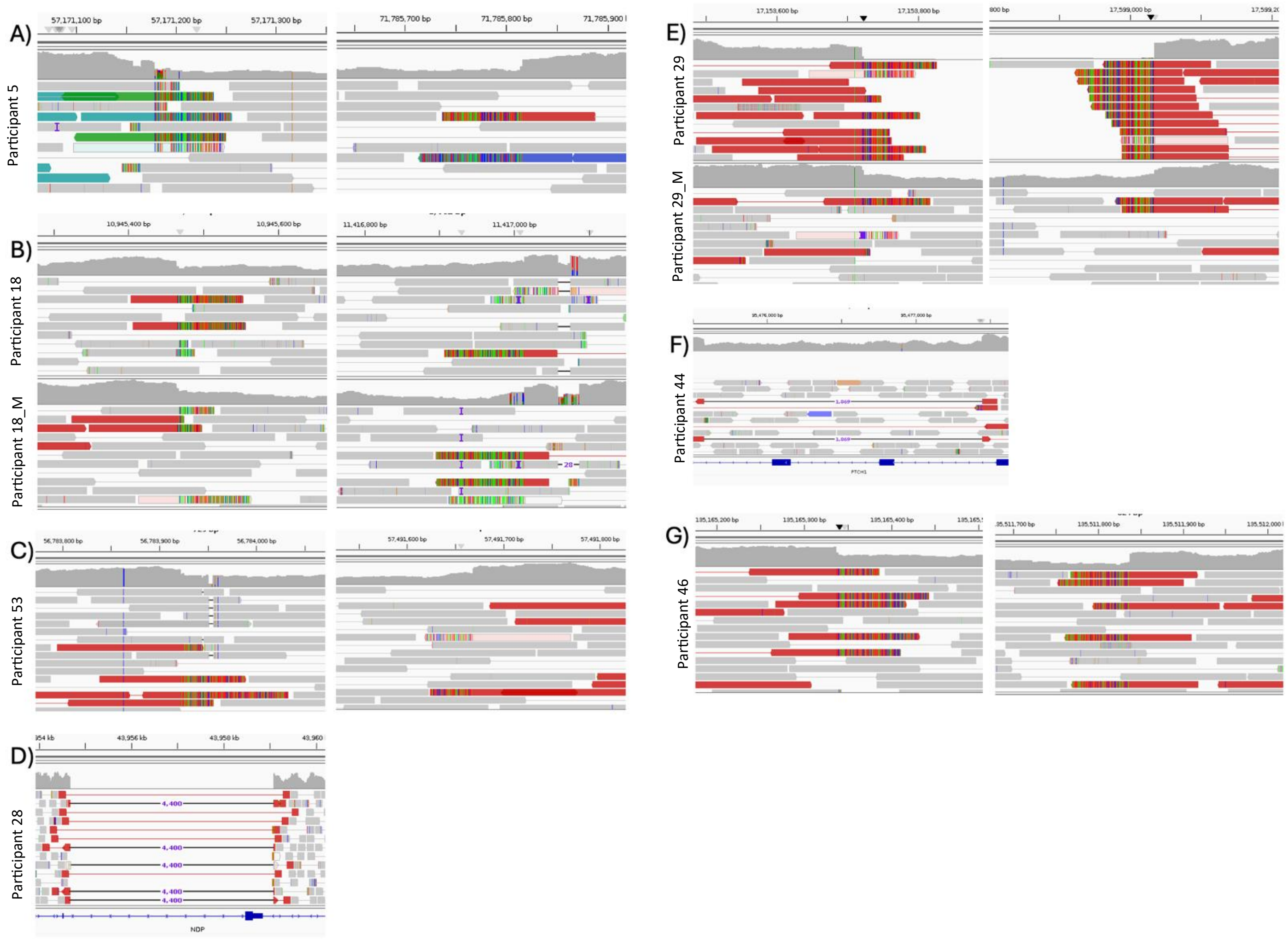


**Supplementary Figure 2.** Deletion breakpoints in A/M participants

A) Participant 5 harbours a heterozygous 14.6 Mb deletion spanning from 10:57171179-71785816, with the breakpoints depicted. B) Participant 18 harbours a heterozygous 471.5 kb deletion spanning from X:10945466-11417014, which was inherited from her heterozygous mother (participant 18_M). C) Participant 53 harbours a heterozygous 707.7 kb deletion spanning from 14:56783923-57491668, with the breakpoints depicted. D) Participant 28 harbours a hemizygous 4.4kb deletion spanning from X:43954691-43959091 within the gene *NDP*. E) Participant 29 harbours a hemizygous 44.5 kb deletion spanning from X:17153721-17599035, which was inherited from his heterozygous mother (participant 29_M). F) Participant 44 harbours a heterozygous 1.8kb deletion spanning from 9:95475579-95477447 within the gene *PTCH1*. G) Participant 46 harbours a heterozygous 346.5kb deletion spanning from 2:135,165,337-135,511,837, with the breakpoints depicted.

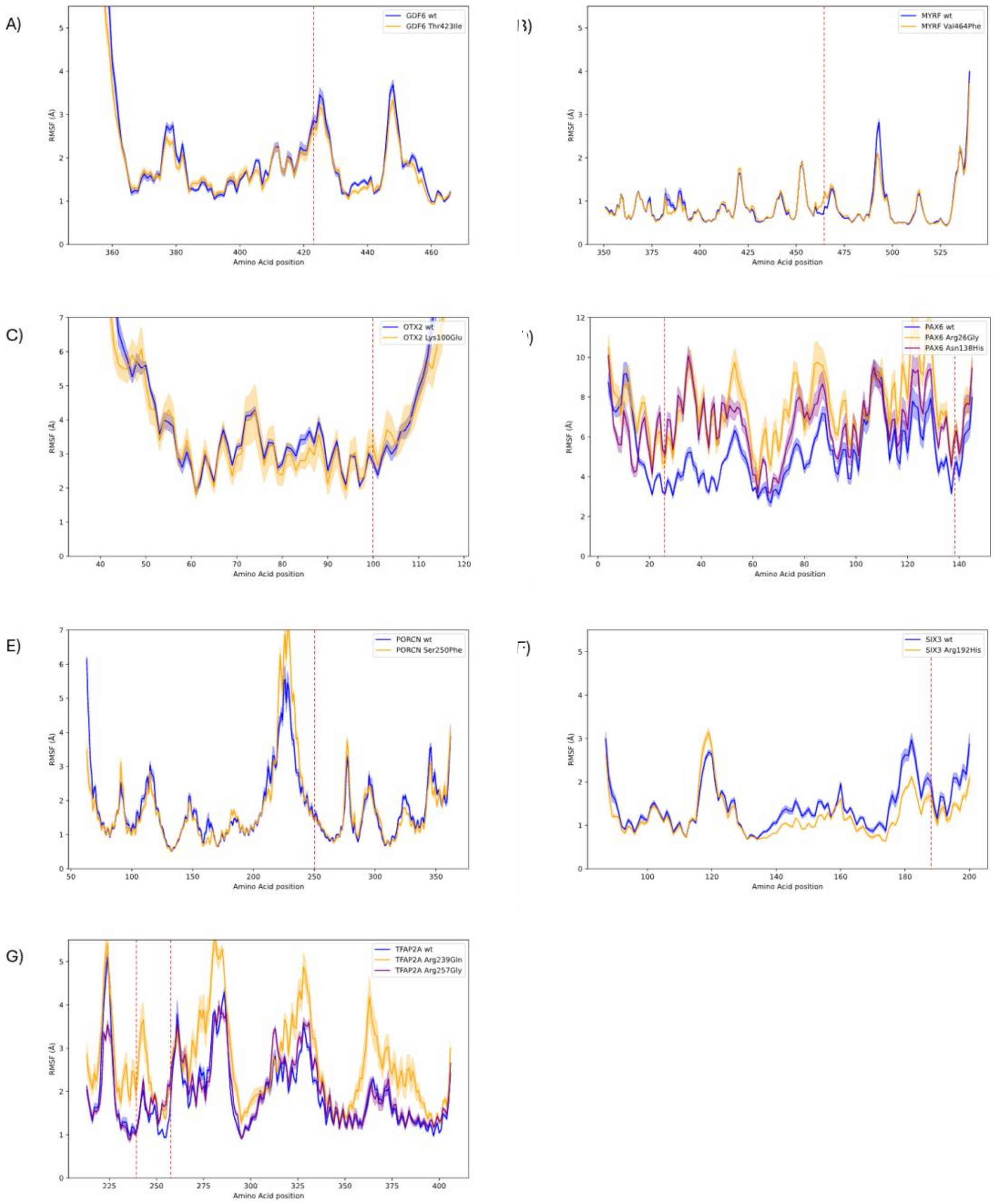


**Supplementary Figure 3.** Individual Root Mean Square Fluctuation for missense variants

Graphs displaying Root mean square fluctuation (RMSF) plots of the Cα atoms for wildtype (wt) and mutant GDF6 (A), MYRF (B), OTX2 (C), PAX6 (D), PORCN (E), SIX3 (F) and TFAP2A (G). The shaded regions represent the standard error of the mean, calculated from four independent replicates. The red dashed line indicates the position of an amino acid substitution.
