## Supplemental Methods for "Refining the genetic landscape of anophthalmia and microphthalmia: a comprehensive framework with deep learning and updated gene panels"

### Supplementary Methods

#### Small variant annotation and filtering

Small variants considered were loss-of-function, missense, and synonymous, intronic and splice variants in the context of splicing. They were annotated with the Ensembl Variant Effect Predictor (VEP, release 99) executed under the everything flag, and the analysis was enriched with four plug-ins: gnomADc, which appends population allele frequencies and constraint metrics from gnomAD v3; MaxEntScan, which reports the predicted change in splice-site strength (Δ); SpliceAI, which provides deep-learning splice-impact delta scores (DS); and UTRannotator, which supplies functional predictions for 5′- and 3′-UTR variants. After annotation, a variant was retained and exported to .tsv only if it met three conditions: (i) rarity, defined as a minor-allele frequency (AF) of ≤0.001 in both gnomAD v3[44] and the Genomics England research cohort; (ii) presence in the proband; and (iii) in-silico evidence of pathogenicity, in which at least one predictor exceeded its threshold (SIFT ≥ 2, PolyPhen-2 ≥ 2, CADD-PHRED ≥ 0, MaxEntScan Δ > 2, or SpliceAI DS > 0.4) or the variant's predicted consequence lay outside the 5′ UTR. Pre-computed AlphaMissense scores were added to the .tsvs post hoc and REVEL scores queried manually.

#### System preparation for molecular dynamics simulations

The most promising candidates for potential destabilizing mutations, (*GDF6*, *LRP2*, *MYRF*, *OTX2*, *PAX6*, *PORCN*, *SIX3*, *TFAP2A*), were modelled using the Colab implementation of AlphaFold2 [1,2]. Each system was truncated to include only the structured regions of the respective proteins. The resulting truncated sequences are provided below under **Sequences**. Mutations were introduced using the CHARMM-GUI PDB Manipulator, which enabled local structural relaxation prior to simulation [3,4]. Protonation states were selected based on a physiological pH of 7. All systems were subsequently parameterized with the AMBER ff14SB force field [5,6] and solvated in a cubic box of TIP3P water molecules [7], ensuring a minimum buffer of 10 Å between the protein surface and the box boundaries. Systems were then neutralized using $Na^+$ and $Cl^-$ ions, and brought to a salt concentration of 150 mM.

#### Molecular Dynamics simulations

The systems underwent energy minimization for up to 5000 steps, beginning with the steepest descent algorithm for the first 2500 steps, followed by the conjugate gradient method. During minimization, weak positional restraints (1 kcal $mol^{-1}$ $Å^{-2}$) were applied to all protein atoms. After minimization, equilibration was carried out under an NVT ensemble for 125 ps. During this phase, the temperature was equilibrated at 300 K. During equilibration, positional restraints were retained, and the simulations used a 1 fs integration time step.

Subsequent production simulations were performed under the NPT ensemble at 1 bar pressure, employing a Langevin thermostat [8] (friction coefficient: 1 $ps^{-1}$) and a Monte Carlo barostat [9]. Periodic boundary conditions and the Particle Mesh Ewald method were used to handle long-range electrostatics, with a real-space cutoff of 9 Å [10]. Van der Waals interactions were truncated at the same real-space cutoff. Protein bonds involving hydrogen atoms were constrained using the SHAKE algorithm, while water molecules were constrained using the SETTLE method, permitting the use of a 2 fs integration time step.

All simulations were performed using the pmemd.cuda module from Amber 2023 [11]. Each system was run in four independent replicates of 100 ns each, resulting in a total aggregated simulation time of 400 ns per system. Root Mean Square Fluctuation (RMSF) analysis was conducted on the backbone Cα atoms using the MDTraj library [12]. Comparative analysis between systems focused on the top 10%

of residues exhibiting the highest fluctuations within each system. All RMSF were normalized based on the mean fluctuations of their respective wild type systems.

**Sequences**

>GDF6_wt

SRHGKRHGKKSRLRCSKKPLHVNFKELGWDDWIIAPLEYEAYHCEGVCDFPLRSHLEPTNHA
IIQTLMNSMDPGSTPPSCCVPTKLTPISILYIDAGNNVVYKQYEDMVVESCGC

>GDF6_Thr423Ile

SRHGKRHGKKSRLRCSKKPLHVNFKELGWDDWIIAPLEYEAYHCEGVCDFPLRSHLEPTNHA
IIQTLMNSMDPGSTPPSCCVPIKLTPISILYIDAGNNVVYKQYEDMVVESCGC

>LRP2_wt

NHHCDSNEWQCANKRCIPESWQCDTFNDCEDNSDEDSSHCAS

>LRP2_Cys3594Arg

NHHRDSNEWQCANKRCIPESWQCDTFNDCEDNSDEDSSHCAS

>OTX_wt

SCPAATPRKQRRERTTFTRAQLDVLEALFAKTRYPDIFMREEVALKINLPESRVQVWFKNRRA
KCRQQQQQQNGGQNKVR

>OTX_Lys100Glu

SCPAATPRKQRRERTTFTRAQLDVLEALFAKTRYPDIFMREEVALKINLPESRVQVWFKNRRA
ECRQQQQQQNGGQNKVR

>MYRF_wt

IKWQPHQQNKWATLYDANYKELPMLTYRVDADKGFNFSVGDDAFVCQKKNHFQVTVYIGM
LGEPKYVKTPEGLKPLDCFYLKLHGVKLEALNQSINIEQSQSDRSKRPFNPVTVNLPPEQVTK
VTVGRLHFSETTANNMRKKGKPNPDQRYFMLVVALQAHAQNQNYTLAAQISERIIVRASNPG
QFESD

>MYRF_VAL464PHE

IKWQPHQQNKWATLYDANYKELPMLTYRVDADKGFNFSVGDDAFVCQKKNHFQVTVYIGM
LGEPKYVKTPEGLKPLDCFYLKLHGVKLEALNQSINIEQSQSDRSKRPFNPVTFNLPPEQVTK
VTVGRLHFSETTANNMRKKGKPNPDQRYFMLVVALQAHAQNQNYTLAAQISERIIVRASNPG
QFESD

>PAX6_wt

SHSGVNQLGGVFVNGRPLPDSTRQKIVELAHSGARPCDISRILQTHADAKVQVLDNQNVSNG
CVSKILGRYYETGSIRPRAIGGSKPRVATPEVVSKIAQYKRECPSIFAWEIRDRLLSEGVCTNDN
IPSVSSINRVLRNLA

>PAX6_Arg26Gly

SHSGVNQLGGVFVNGRPLPDSTGQKIVELAHSGARPCDISRILQTHADAKVQVLDNQNVSNG
CVSKILGRYYETGSIRPRAIGGSKPRVATPEVVSKIAQYKRECPSIFAWEIRDRLLSEGVCTNDN
IPSVSSINRVLRNLA

>PAX6_Asn138His

SHSGVNQLGGVFVNGRPLPDSTRQKIVELAHSGARPCDISRILQTHADAKVQVLDNQNVSNG
CVSKILGRYYETGSIRPRAIGGSKPRVATPEVVSKIAQYKRECPSIFAWEIRDRLLSEGVCTNDN
IPSVSSIHRVLRNLA

>PORCN_wt

SLYHFFQLHMVWVVLLSLLCYLVLFLCRHSSHRGVFLSVTILIYLLMGEMHMVDTVTWHKM
RGAQMIVAMKAVSLGFDLDRGEVGTVPSPVEFMGYLYFVGTIVFGPWISFHSYLQAVQGRPL
SCRWLQKVARSLALALLCLVLSTCVGPYLFPYFIPLNGDRLLRNKKRKARGTMVRWLRAYES
AVSFHFSNYFVGFLSEATATLAGAGFTEEKDHLEWDLTVSKPLNVELPRSMVEVVTSWNLPM
SYWLNNYVFKNALRLGTFSAVLVTYAASALLHGFSFHLAAVLLSLAFITYVEH

>PORCN_Ser250Phe

SLYHFFQLHMVWVVLLSLLCYLVLFLCRHSSHRGVFLSVTILIYLLMGEMHMVDTVTWHKM
RGAQMIVAMKAVSLGFDLDRGEVGTVPSPVEFMGYLYFVGTIVFGPWISFHSYLQAVQGRPL
SCRWLQKVARSLALALLCLVLSTCVGPYLFPYFIPLNGDRLLRNKKRKARGTMVRWLRAYES
AVFFHFSNYFVGFLSEATATLAGAGFTEEKDHLEWDLTVSKPLNVELPRSMVEVVTSWNLPM
SYWLNNYVFKNALRLGTFSAVLVTYAASALLHGFSFHLAAVLLSLAFITYVEH

>SIX3_wt

FSPEQVASVCETLEETGDIERLGRFLWSLPVAPGACEAINKHESILRARAVVAFHTGNFRDLYHI
LENHKFTKESHGKLQAMWLEAHYQEAEKLRGRPLGPVDKYRVRKKFPLP

>SIX3_Arg192His

FSPEQVASVCETLEETGDIERLGRFLWSLPVAPGACEAINKHESILRARAVVAFHTGNFRDLYHI
LENHKFTKESHGKLQAMWLEAHYQEAEKLRGRPLGPVDKYHVRKKFPLP

>TFAP2A_wt

FCSVPGRLSLLSSTSKYKVTVAEVQRRLSPPECLNASLLGGVLRRAKSKNGGRSLREKLDKIG
LNLPAGRRKAANVTLLTSLVEGEAVHLARDFGYVCETEFPAKAVAEFLNRQHSDPNEQVTRK
NMLLATKQICKEFTDLLAQDRSPLGNSRPNPILEPGIQSCLTHFNLISHGFGSPAVCAAVTALQN
YLTE

>TFAP2A_Arg239Gln

FCSVPGRLSLLSSTSKYKVTVAEVQQRLSPPECLNASLLGGVLRRAKSKNGGRSLREKLDKIG
LNLPAGRRKAANVTLLTSLVEGEAVHLARDFGYVCETEFPAKAVAEFLNRQHSDPNEQVTRK
NMLLATKQICKEFTDLLAQDRSPLGNSRPNPILEPGIQSCLTHFNLISHGFGSPAVCAAVTALQN
YLTE

>TFAP2A_ARG257Gly

FCSVPGRLSLLSSTSKYKVTVAEVQRRLSPPECLNASLLGGVLGRAKSKNGGRSLREKLDKIG
LNLPAGRRKAANVTLLTSLVEGEAVHLARDFGYVCETEFPAKAVAEFLNRQHSDPNEQVTRK
NMLLATKQICKEFTDLLAQDRSPLGNSRPNPILEPGIQSCLTHFNLISHGFGSPAVCAAVTALQN
YLTE

### References


1. Jumper, John, et al. "Highly accurate protein structure prediction with AlphaFold." *nature* 596.7873 (2021): 583-589.
2. Mirdita, Milot, et al. "ColabFold: making protein folding accessible to all." Nature methods 19.6 (2022): 679-682.
3. Jo, Sunhwan, et al. "CHARMM-GUI: a web-based graphical user interface for CHARMM." Journal of computational chemistry 29.11 (2008): 1859-1865.
4. Park, Sang-Jun, et al. "CHARMM-GUI PDB manipulator: various PDB structural modifications for biomolecular modeling and simulation." Journal of molecular biology 435.14 (2023): 167995.
5. Maier, James A., et al. "ff14SB: improving the accuracy of protein side chain and backbone parameters from ff99SB." Journal of chemical theory and computation 11.8 (2015): 3696-3713.
6. Brooks, Bernard R., et al. "CHARMM: the biomolecular simulation program." Journal of computational chemistry 30.10 (2009): 1545-1614.
7. Jorgensen, William L., et al. "Comparison of simple potential functions for simulating liquid water." The Journal of chemical physics 79.2 (1983): 926-935.
8. Loncharich, Richard J., Bernard R. Brooks, and Richard W. Pastor. "Langevin dynamics of peptides: The frictional dependence of isomerization rates of N-acetylalanyl-N′-methylamide." Biopolymers: Original Research on Biomolecules 32.5 (1992): 523-535.
9. Åqvist, Johan, et al. "Molecular dynamics simulations of water and biomolecules with a Monte Carlo constant pressure algorithm." Chemical physics letters 384.4-6 (2004): 288-294.
10. Darden, Tom, Darrin York, and Lee Pedersen. "Particle mesh Ewald: An N log (N) method for Ewald sums in large systems." Journal of chemical physics 98 (1993): 10089-10089.
11. Case, David A., et al. Amber 2023. University of California, San Francisco, 2023.
12. McGibbon, Robert T., et al. "MDTraj: a modern open library for the analysis of molecular dynamics trajectories." Biophysical journal 109.8 (2015): 1528-1532.
